# Spatial AI quantification of inflammatory and architectural features differentiates pathologist-defined active ulcerative colitis from histological remission

**DOI:** 10.64898/2026.01.27.26344949

**Authors:** Dylan Windell, Alastair Magness, Rongqi Li, Tom Davis, Helena Thomaides Brears, Sarah Larkin, Cayden Beyer, Paul Aljabar, Reema Kainth, Phil Wakefield, Caitlin Langford, Nicholas Powell, Mark Delegge, Adrian C. Bateman, Roger Feakins, Eve Fryer, Robert Goldin, Jonathan Landy

## Abstract

**Background and Aims:** Artificial intelligence (AI) is applied to histological assessment in inflammatory bowel disease (IBD), but most approaches quantify features without considering anatomical location. We developed and evaluated PAIR-IBD (Perspectum AI Reading in IBD), an AI system quantifying inflammatory cell, crypt injury, and epithelial damage within mucosal compartments to distinguish histologically-defined active disease, remission, and inconclusive cases in ulcerative colitis (UC), in a single clinical network.

**Methods:** A deep-learning ensemble was trained on three IBD datasets to identify lymphocytes, neutrophils, eosinophils, and plasma cells, and segment crypts, lamina propria (LP), surface epithelium and muscularis mucosae. Inflammatory cell densities and crypt injury metrics (mucin depletion, solidity, roughness, branching, and abscesses) were quantified. PAIR-IBD outputs were compared between pathologist-defined active UC and histological remission, evaluated in discordant cases and correlated with manual grading.

**Results:** Compared with Nancy remission, active biopsies showed 2.1-fold higher LP neutrophil, 5.3-fold higher crypt neutrophil and 1.2-fold higher LP lymphocyte density. Active UC showed increased mucin depletion, crypt branching, and crypt abscesses, with reduced crypt solidity (p<0.0001 for all). PAIR-IBD metrics correlated with manual pathology scores and global indices, strongest for mucin depletion, crypt neutrophils, crypt roughness and reduced crypt solidity (r_s_=0.50–0.73). Up to 89% of inconclusive biopsies aligned with histological remission-like profiles for individual metrics, particularly mucin depletion.

**Conclusion:** PAIR-IBD provides spatially resolved, quantitative assessment of inflammation, epithelial injury and crypt architecture in UC, with potential to support the characterization of inconclusive histology, complement scoring and contribute to histological endpoint assessment in clinical trials, pending validation.

**What is already known on this topic:** Histological remission is a key treatment target in ulcerative colitis (UC), but assessment relies on ordinal pathologist scoring systems (Nancy Index, Robarts Histopathological Index, Geboes Score) that are subject to inter-rater variability.

**What this study adds:** We developed and evaluated PAIR-IBD, an artificial intelligence system quantifying inflammatory cell populations, crypt injury and epithelial damage across mucosal compartments in a single clinical network cohort. PAIR-IBD distinguished pathologist-defined active UC from remission and showed that inconclusive biopsies display a mixed rather than uniformly active or remission-like histological profile.

**How this study might affect research, practice or policy:** These findings support further evaluation of PAIR-IBD as an adjunct to conventional histological scoring, with potential applications in clinical trial endpoint assessment and adjudication of discordant biopsies, pending external validation.

## INTRODUCTION

Inflammatory Bowel Disease (IBD) is a chronic inflammatory disease with effective therapies but no cure to date. IBD affects approximately 721 per 100,000 people in the US^1^ and 781 per 100,000 people in the UK^2,3^ and is increasing globally^4,5^. The main subtypes, Crohn’s Disease (CD) and Ulcerative Colitis (UC), cause debilitating symptoms and carry an increased risk of colorectal cancer.

As highlighted by ECCO^6^, UC features include immune cell infiltrations, crypt injury and epithelial damage across the colon. The most severe histologic features are epithelial damage, erosion or ulceration^7^ while neutrophils are considered the defining feature of active UC, appearing in the lamina propria (LP), crypt epithelium (cryptitis), or crypt lumens (crypt abscesses).

Histologic remission is associated with long-term clinical remission and a reduction in cancer risk^8,9^. Histology-based assessment can detect residual microscopic inflammation that may persist in up to 40% of adult with UC exhibiting endoscopic healing^10^. Acute inflammatory indicators from histopathology are associated with a two- to three-fold increased risk of colitis relapse^11^, and basal plasmacytosis predicts UC clinical relapse in adults with UC with complete mucosal healing^12^. Histological remission is therefore an important therapeutic goal in UC clinical trials^13–15^ alongside endoscopic remission.

The most widely used histological indices in clinical trials are the Nancy Index (NI), Robarts Histopathological Index (RHI) and the Geboes score (GS). These indices incorporate different combinations of inflammatory, epithelial and architectural disease features^16–20^. Even among expert GI pathologists, inter-rater reliability remains only moderate for GS and NI, while RHI requires further validation^11,18,21–23^. Differing criteria across the >30 UC histological indices contribute to variable inter-rater reliability and divergent disease classification^11^.

Multiple artificial intelligence (AI) models for histological analysis have been designed for UC^23,24^. The AI systems quantify various cell populations and crypt features^25–34^ but do not jointly assess inflammatory and crypt morphological features across the spectrum of UC severity or account for how these features are distributed through biopsy depth from the surface epithelium to the deeper mucosa. Accordingly, there is a need for a single quantitative support tool for pathologists that moves beyond coarse ordinal indices to accurately quantify both inflammatory indicators and architectural changes in a spatially-resolved manner^35^. In this study we sought to develop and evaluate PAIR-IBD, a comprehensive AI tool that jointly quantifies regional inflammatory cell populations, crypt injury and epithelial damage in UC. We evaluated whether PAIR-IBD enabled 1) clear differentiation between histologically active and UC in histological remission across 3 histological indices (NI, RHI, and SGS), and 2) positioned inconclusive scored biopsies along a spectrum of mucosal injury and healing.

## MATERIALS AND METHODS

### Study Design and Populations

The West Hertfordshire IBD Technology Study (WHITS) is a prospective, longitudinal, observational, non-interventional study of individuals living with IBD (NCT05000242)^36^. We applied PAIR-IBD (*v2026-07*) to baseline UC histology from WHITS participants recruited between 2021 and 2023 (**Supplementary Figure 1**). Inclusion criteria were digitised datasets above a quality-controlled threshold for biopsy staining and clinical confirmation of UC diagnosis (performed independently of this work) (**Supplementary Methods**).

### Ethics and study consent

Informed consent was obtained from all study participants for the WHITS trial, including the use of biopsy material and associated clinical data for digital image analysis and secondary research. Ethical approval was provided by the London Queen Square research ethics committee (ethics reference 20/LO/0349).

### Data collection

#### Endoscopy, blood tests, stool and biopsy sample collection

All individuals with UC attended a baseline assessment at West Hertfordshire Hospitals NHS Trust, including blood tests, stool, medical history, treatment and anthropometric data. Endoscopic assessment was performed as part of standard clinical care, and where available, endoscopic activity was scored by a single expert endoscopist using the Mayo Endoscopic Score (MES). Mucosal biopsies were obtained during endoscopy for histological assessment, with an average of 3 biopsies per subject.

#### Biopsy digitization and selection

Whole slide images (WSIs) were digitised at 40× on two scanners and analysed on the biopsy section with the highest lymphocyte density (**Figure 1**; **Supplementary Methods**).

**Figure 1.**
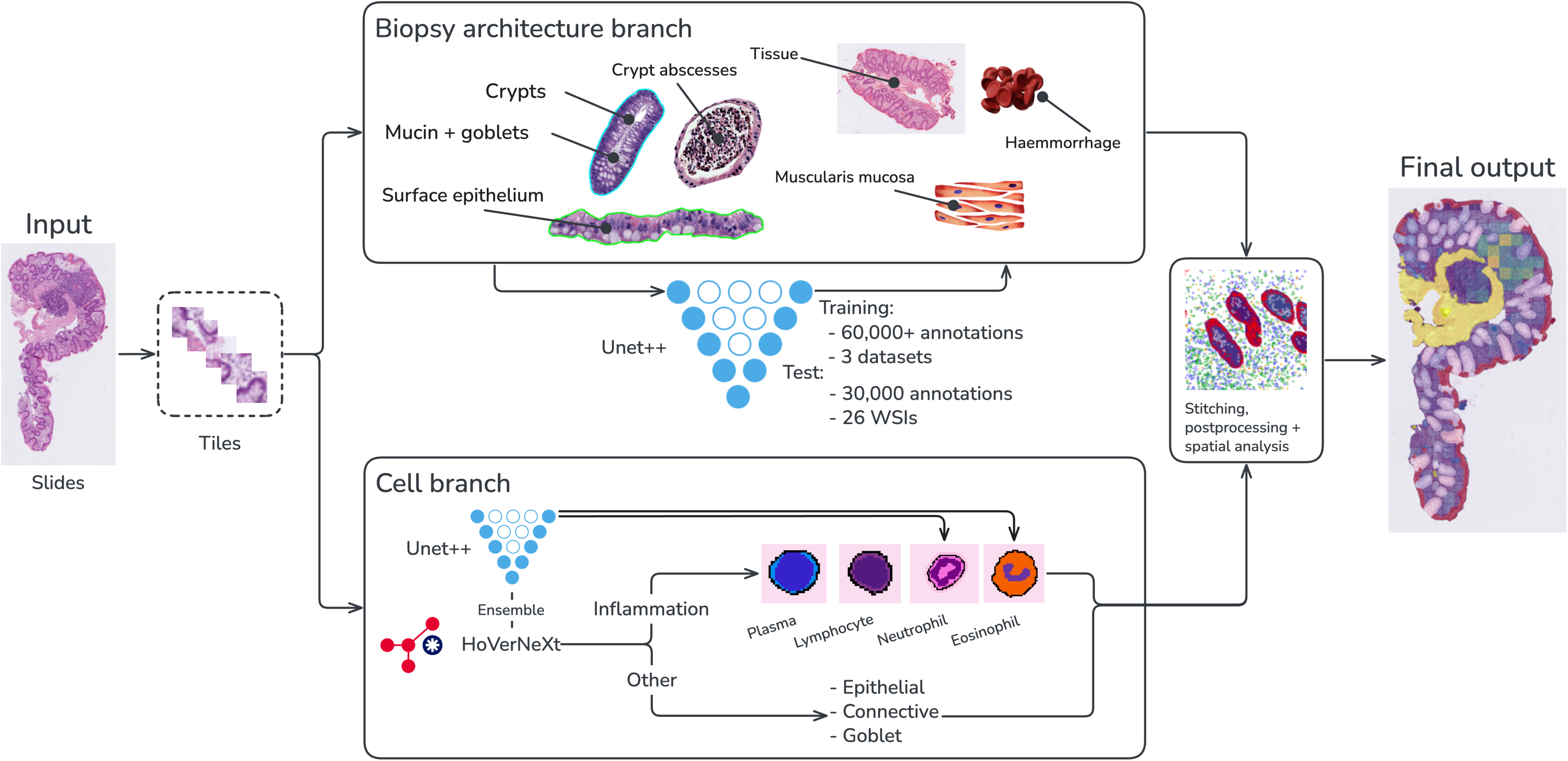
Integrated tissue segmentation and cell classification enables quantitation of IBD phenotypes and highlighting of features in whole slides for pathologist decision support.

#### Histological scoring

WSIs were scored independently by two pathologists with 14 and 40+ years’ experience, respectively, blinded to clinical data. All scoring was performed using a universal template (**Supplementary Table 1**), enabling subsequent conversion to the NI, SGS and RHI scores.

Histological remission cut-offs were NI≤0, SGS≤2A.1 and RHI≤3; discordant reads were classed as inconclusive^37–39^.

#### Development of PAIR-IBD for feature detection in IBD

PAIR-IBD is an ensemble of convolutional neural networks that segments key mucosal compartments and classifies inflammatory cell types on Hematoxylin & Eosin (H&E)-stained whole-slide images. The system quantifies inflammatory cell densities and crypt-level injury features within defined mucosal regions. Model performance was assessed on 30,000 held-out image-annotation pairs from 26 WSIs, with slide-level separation between training and testing datasets (**Supplementary Methods**). Annotations were performed by a single specialist gastrointestinal pathologist, who annotated the full set of images used for training and evaluation; an engineer supported image preparation and annotation tooling. Annotation was sparse, with a defined subset of cells per class labelled rather than exhaustive annotation of every instance within each image.

#### Quantification of inflammatory cell, crypt injury and epithelial damage with PAIR-IBD

AI-based quantification was performed on the biopsy section with the highest lymphocyte density (**Figure 1**). Histological analyses were performed at the biopsy (WSI) level rather than at the subject level. This was a retrospective analysis of prospectively acquired histological samples.

Five inflammatory metrics were derived:

1) LP lymphocyte density (cell/mm^2^); 2) neutrophil density (cell/mm^2^) in the i) LP and ii) surface epithelium iii) crypt epithelium; 3) LP eosinophil density (cell/mm^2^); 4) LP plasma cell density (cell/mm^2^); and 5) combined plasma cell and lymphocyte density (cell/mm^2^) in five 100 μm zones from the surface epithelium (SE) towards the crypt bases and muscularis mucosa (MM).

In addition, the following 5 metrics of crypt injury and epithelial damage were derived:

1) mucin depletion, defined as loss of intraluminal mucin relative to total crypt area; 2) crypt abscess fraction, defined as the proportion of crypts containing intraluminal inflammatory cells; 3) crypt solidity and 4) crypt roughness, which captured deviations from normal crypt shape using area- and perimeter-based measures relative to the crypt convex hull; 5) crypt branching, a quantification of abnormal crypt architecture based on enumeration of branches within the crypt skeleton. All metrics were averaged across crypts within each analysed tissue section. Detailed definitions and equations are provided in the **Supplementary Methods**.

### Statistical analyses

Cohort characteristics were summarised descriptively. Previously published neutrophil thresholds of 30.4 cells/mm² for the lamina propria and 10.1 cells/mm² for the crypt epithelium were applied as originally reported^25^, without cohort-specific optimisation. Exploratory cohort-specific thresholds were subsequently derived by maximising the Youden index. Sensitivity, specificity, accuracy, positive predictive value and negative predictive value were calculated against consensus NI, SGS, and RHI classifications. A sensitivity analysis further restricted consensus RHI remission biopsies to those in which both pathologists scored the LP and crypt neutrophil components as zero. Differences between AI-derived metrics across disease states were assessed using Wilcoxon rank-sum test, and associations with manual pathology scoring were assessed using Spearman’s rank correlation (r_s_). Inter-rater reliability was summarised using overall percentage agreement (OPA), weighted Cohen’s kappa and intraclass correlation (ICC) for RHI. P values <0.05 were considered statistically significant. Benjamini–Hochberg adjustment was used for group comparisons and Bonferroni adjustment for multiple comparisons.

## RESULTS

### Study populations

Data from 83 individuals with confirmed UC and 272 digitised WSI of sufficient quality, recruited across two sites within the West Hertfordshire Hospitals NHS Trust (2021-2023) were included **(Supplementary Figure 1)**. Mean age was 43 years and 51% were male (**Table 1**). Baseline biomarkers suggested an overall active cohort, with elevated mean CRP and F-calprotectin, and low-normal albumin and haemoglobin. Most subjects were receiving treatment, predominantly topical therapy or biologic/small-molecule therapy. Eighteen WSIs were excluded from the original 290; ten as non-colonic or other stains, two for poor quality, two by quality-control failure and four through pipeline failure.

**Table 1:**
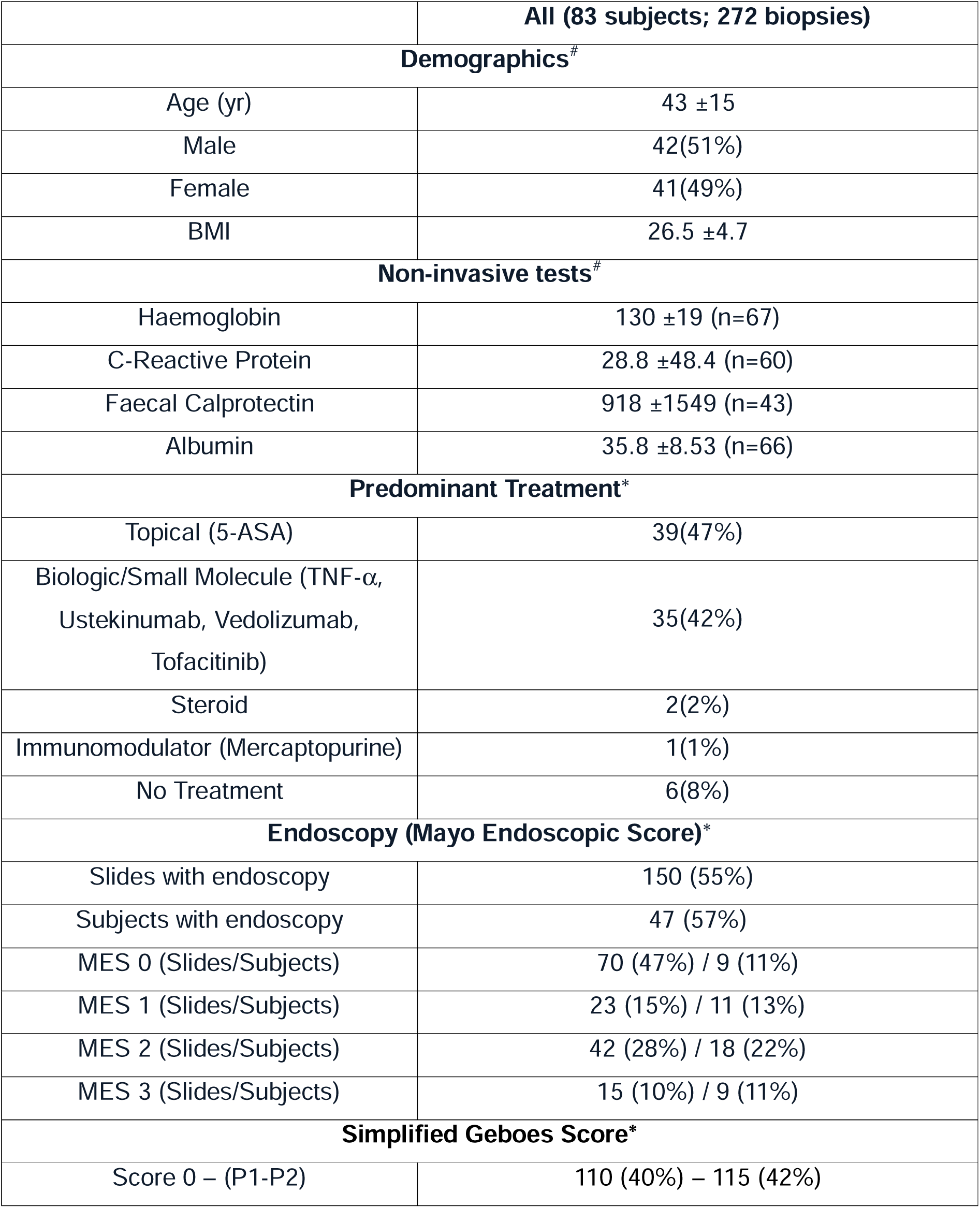
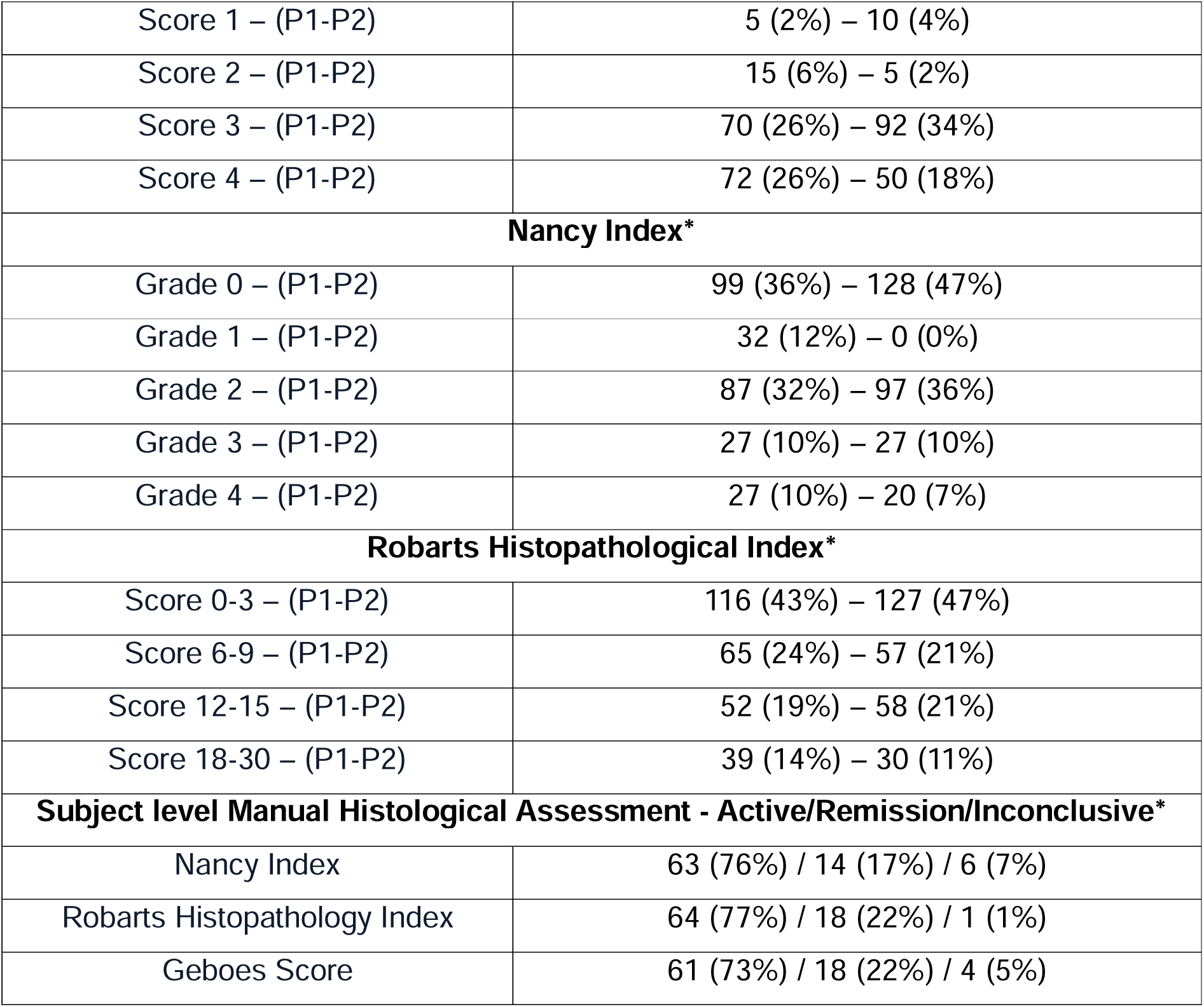
Study populations. Low κ values for certain features reflect prevalence effects due to the predominance of score 0 rather than true inter-reader disagreement, a recognised limitation of κ in highly imbalanced datasets^55^.

Endoscopic scores were available for 150 of 272 biopsies (55%), representing 47 of 83 subjects (57%). Overall, 62% of slides and 43% of subjects were in endoscopic remission, as per the MES 0-1 classification.

### Scoring system agreement

Agreement between pathologists across the universal template variables ranged from 65% to 84%. Final converted indices showed moderate to strong agreement, with NI agreement of 71% (κ=0.53), SGS agreement of 51% (κ=0.58), and RHI ICC of 0.86. At the subject level (n=83), using the worst-biopsy approach, active/remission/inconclusive distributions were 76%/17%/7% for NI, 77%/22%/1% for RHI and 73%/22%/5% for SGS. Full score breakdowns are shown in Table 1 and Supplementary Table 2.

MES also showed strong alignment with manual histology, with agreement across individual readers, consensus reads and all three histological indices ranging from 83% to 93%.

### AI model performance

PAIR-IBD model performance was assessed on 26 held-out, annotated WSIs sampling the IBD disease spectrum. Tile-based segmentation assessment across six tissue classes showed near-perfect area overlap for tissue foreground and strong performance for lamina propria and crypts, with lower performance for surface epithelium and muscularis mucosa, the latter most likely reflecting orientation and sectioning variability. Instance-level evaluation of crypts demonstrated balanced detection and high segmentation accuracy (**Supplementary Table 3-4**). Cell-level detection against pathologist review was similarly high overall (class-agnostic precision 0.998, recall 0.934, F1 0.965, 95% CI 0.945–0.980), driven by near-complete lymphocyte detection (recall 0.992) alongside lower recall for eosinophils, neutrophils and plasma cells (recall 0.756–0.779), while precision remained ≥0.989 across all four cell types, indicating that missed detections rather than cross-class misclassification accounted for most errors (Supplementary Table 5).

Nine of 11 PAIR-IBD metrics correlated with their corresponding pathologist feature scores for both pathologists, with eosinophil density showing a weak associated for one pathologist only, while plasma-cell density showed no significant association for either pathologist. The strongest associates were observed for crypt neutrophils (rs=0.59–0.67) and both crypt injury and epithelial damage features including mucin depletion (rs=0.68–0.73), reduced crypt solidity (rs=0.62–0.66), and crypt roughness (rs=0.62–0.65) (**Supplementary Figure 2-6**). Across NI, RHI and SGS, the strongest correlations were observed for mucin depletion (rs=0.65–0.69), crypt neutrophils (rs=0.59–0.67), crypt roughness (rs=0.52–0.63) and reduced crypt solidity (rs=0.50– 0.61), while LP neutrophil correlations were moderate (rs=0.45–0.49) (**Figure 2**).

**Figure 2.**
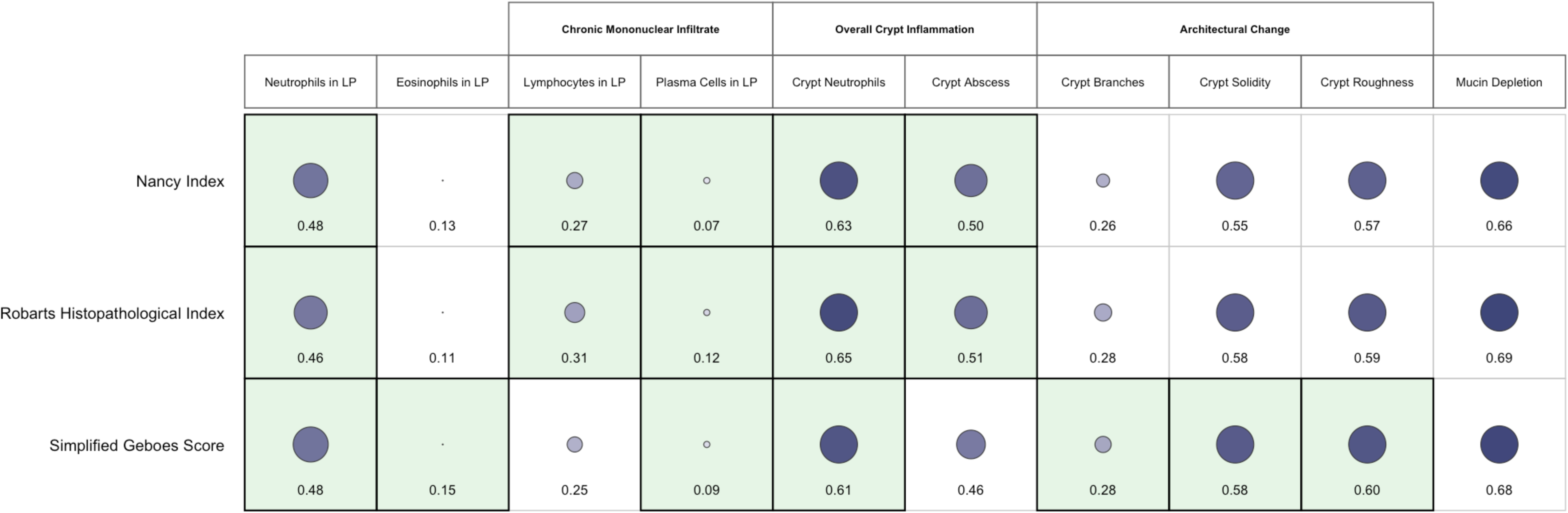
Correlation map showing individual AI-derived histologic features related to the three major IBD activity scoring systems: Nancy Index, Robarts Histopathological Index, and Simplified Geboes Score. Larger, darker bubbles indicate stronger correlations between each AI feature and the corresponding pathologist rating. Features are grouped into clinically relevant domains, including chronic inflammatory infiltrates, crypt-related inflammation, and architectural distortion. Highlighted cells denote the specific tissue features that are formally assessed within each scoring system itself (e.g., neutrophils and eosinophils in the Geboes Score; chronic mononuclear infiltrate and plasma cells in the Nancy and Robarts indices; crypt neutrophils and abscesses in systems that evaluate crypt inflammation).

### Characterisation of active UC with PAIR-IBD

Active UC showed increased inflammatory cell burden in both the LP and crypts. Compared with histological remission, LP neutrophil density was higher in active disease (101.67 vs 49.54 cells/mm²; mean difference [MD] 52.12, 95% CI 37.42– 67.65; **Figure 3A**), with 92% of active biopsies exceeding the published 30.4 cells/mm² threshold^25^. Both surface-epithelium neutrophil density (67.18 vs 36.67 cells/mm²; MD 30.51, 95% CI 17.99–43.19) and crypt neutrophil density were markedly higher in active UC (57.90 vs 10.92 cells/mm²; MD 46.98, 95% CI 35.58– 60.18; **Figure 3B**). Moreover, LP lymphocyte density (2490.19 vs 2021.59 cells/mm²; MD 468.59, 95% CI 226.07–719.97) were elevated in active disease (**Figure 4A**). Both LP eosinophils and LP plasma cell density did not differ significantly. Active disease also showed greater mucin depletion (0.85 vs 0.70; MD 0.15, 95% CI 0.12–0.17; **Figure 5A**), crypt abscesses (0.05 vs 0.01; MD 0.04, 95% CI 0.02–0.06; **Figure 5B**), crypt branching (1.56 vs 1.04; MD 0.52, 95% CI 0.34–0.71; **Figure 6A**), and crypt roughness (1.18 vs 1.08; MD 0.10, 95% CI 0.08–0.12), together with reduced crypt solidity (0.85 vs 0.92; MD −0.07, 95% CI −0.08 to −0.06; **Figure 6B**; all p<0.0001). Index-specific metrics, including SDs, are shown in **Supplementary Table 6-8**. Among NI-derived metrics, the strongest discrimination of histological active versus remission disease was observed for mucin depletion (AUC 0.88, 95% CI 0.84–0.92), crypt roughness (AUC 0.88, 95% CI 0.83–0.92), crypt neutrophils (AUC 0.86, 95% CI 0.81–0.90) and reduced crypt solidity (inverted AUC 0.85, 95% CI 0.80–0.90), followed by LP neutrophils (AUC 0.78, 95% CI 0.72–0.83), crypt abscesses (AUC 0.73, 95% CI 0.67–0.78), crypt branching (AUC 0.72, 95% CI 0.66–0.78), lymphocyte density (AUC 0.67, 95% CI 0.60–0.73) and surface-epithelium neutrophils (AUC 0.66, 95% CI 0.59–0.73).

**Figure 3.**
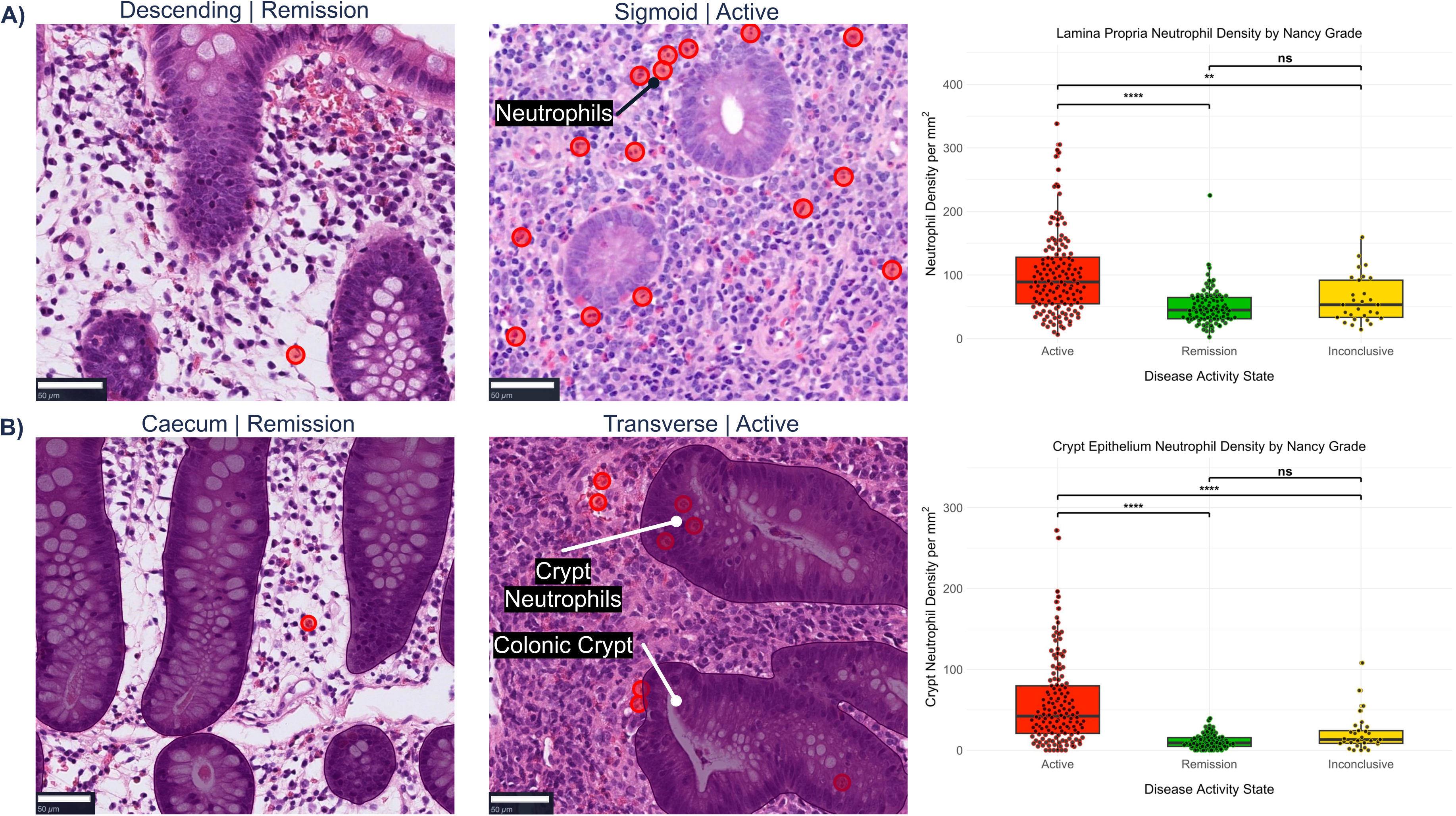
Representative AI-generated overlays showing neutrophil changes in UC and their comparison with two pathologist scores. (A) Spatial distribution of neutrophils within the lamina propria and (B) neutrophils within the crypts visualized in remission and active UC. Box-and-whisker plots show corresponding quantitative AI measurements for neutrophil and neutrophil crypt metrics within manual histological consensus scoring: Nancy Index remission (≤0), active disease (≥1), and inconclusive (non-agreed) cases.

**Figure 4.**
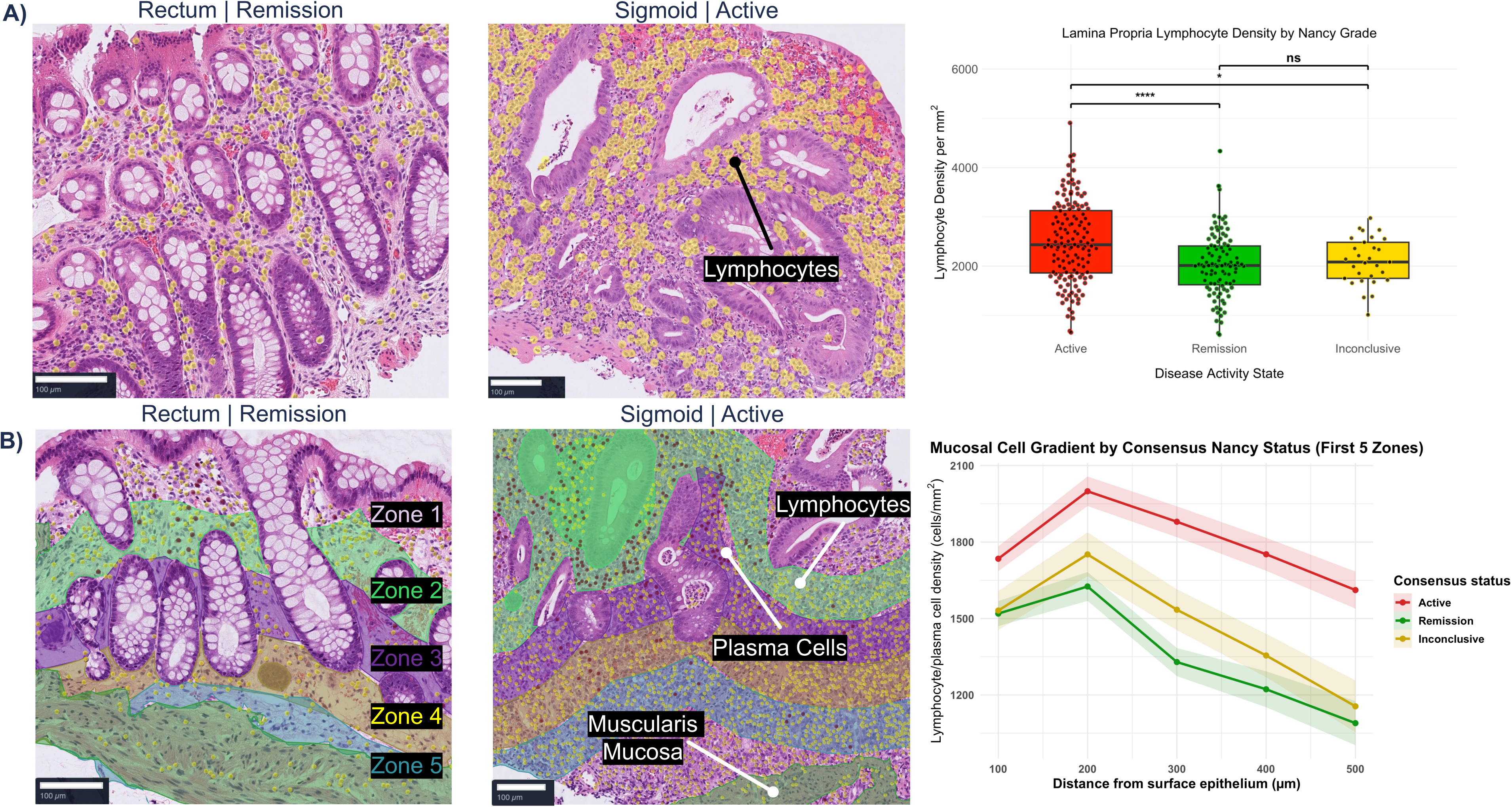
Representative AI-generated overlays showing plasma cell and lymphocyte changes in UC and their comparison with two pathologist scores. (A) Regional distribution of both plasma cell and lymphocytes in zonal regions from the surface epithelium to the muscularis mucosa. Densities of plasma cells and lymphocytes were measured across the lamina propria within each 100 μm zone. (B) Lymphocyte densities within the lamina propria visualised in remission and active UC. Box-and-whisker plots show corresponding quantitative AI measurements for plasma cells and lymphocytes within manual histological consensus scoring: Nancy Index remission (≤0), active disease (≥1), and inconclusive (non-agreed) cases.

**Figure 5.**
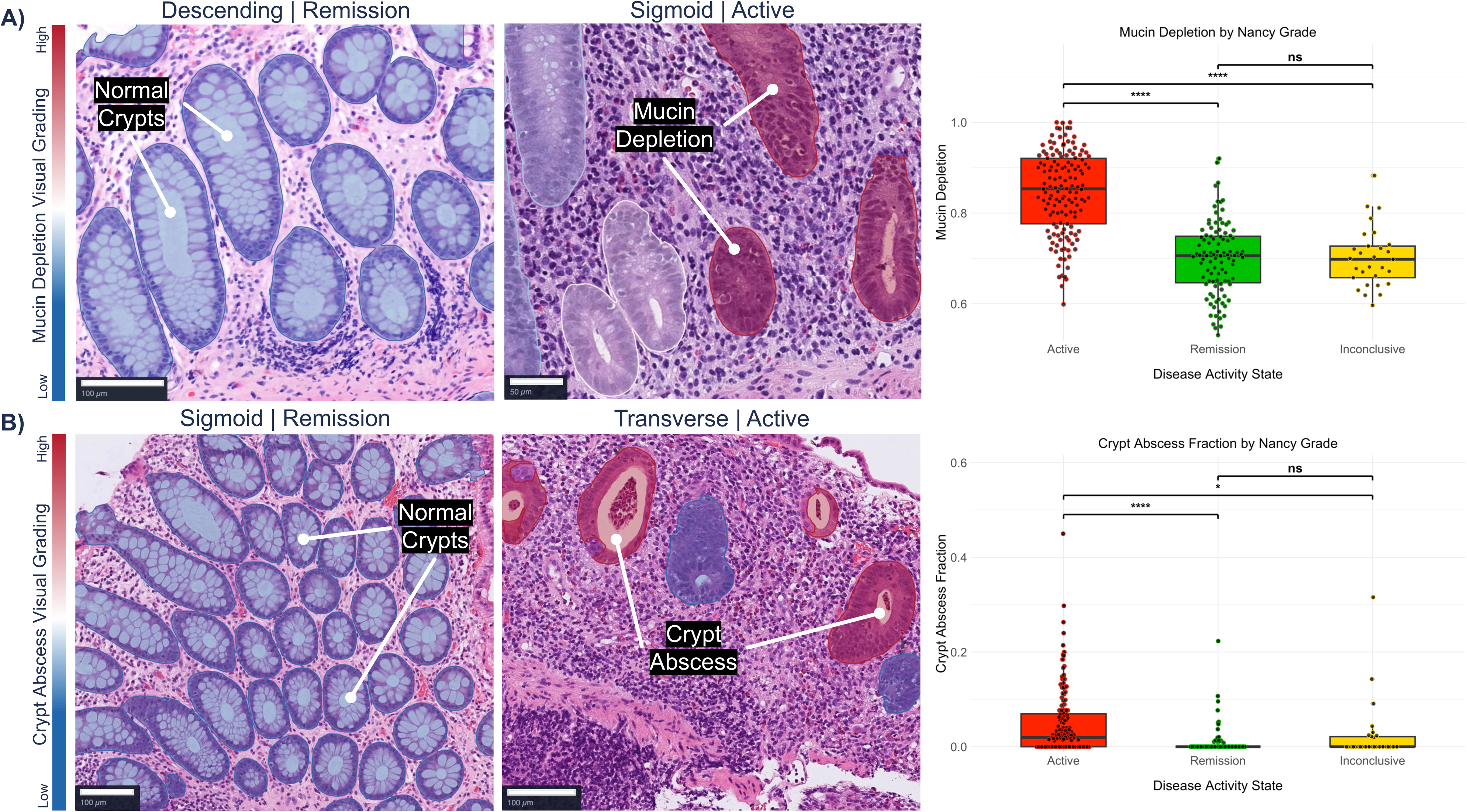
Representative AI-generated overlays showing crypt morphology metric changes in UC and their comparison with two pathologist scores. (A) Mucin depletion is displayed as a loss of mucin content across crypts and in remission and active UC. (B) Crypt abscess presence is highlighted in active disease cases. Box-and-whisker plots show corresponding quantitative AI measurements for mucin depletion and crypt abscess fraction within manual histological consensus scoring: Nancy Index remission (≤0), active disease (≥1), and inconclusive (non-agreed) cases. Visual scale showing increase disease activity in red and lower disease activity in blue.

**Figure 6.**
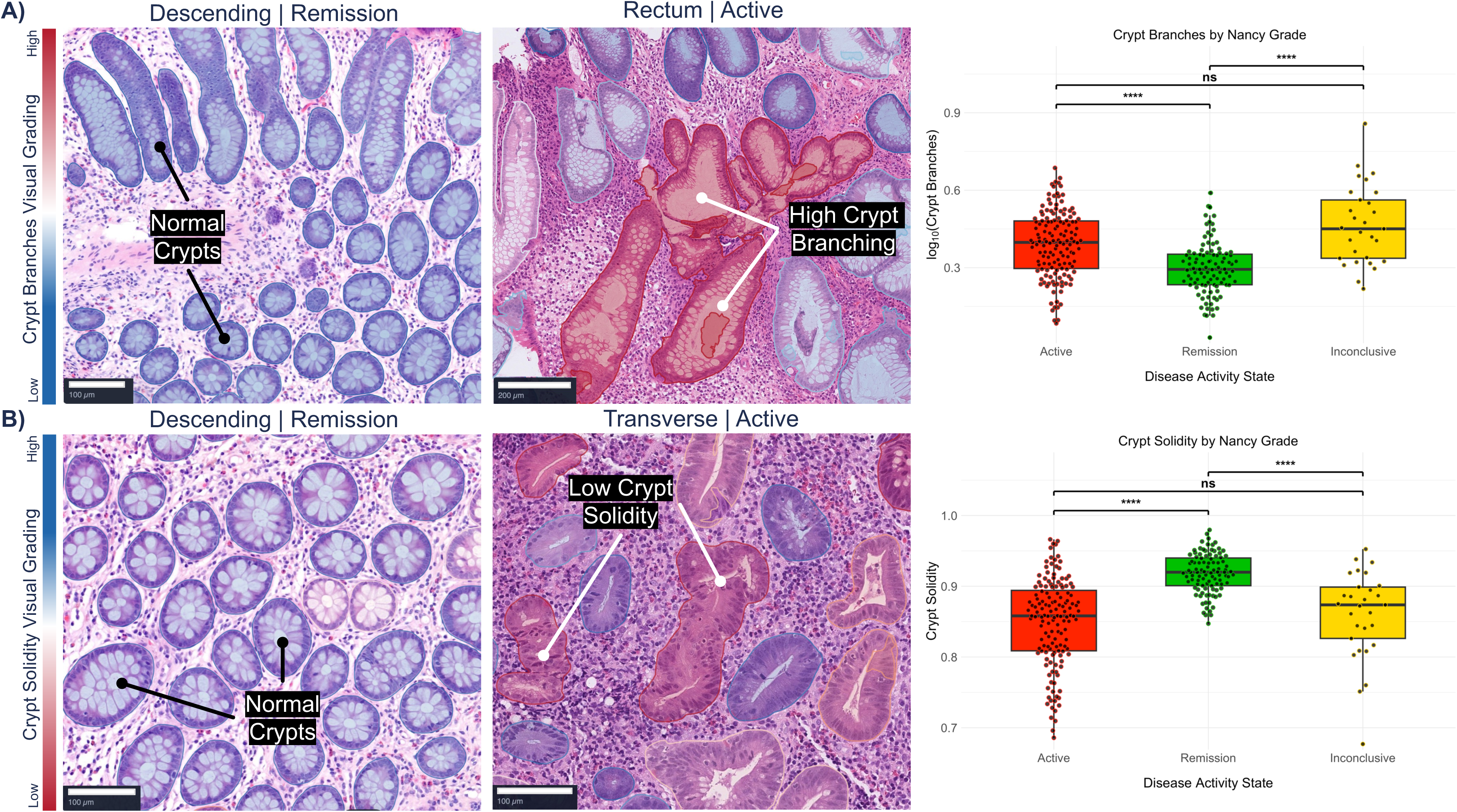
Representative AI-generated overlays showing crypt morphology metric changes in UC and their comparison with two pathologist scores. (A) Crypt branching is represented across groups of branched crypts in active disease cases. (B) Crypt solidity is displayed as whether crypt shape is deformed or altered in active UC with low crypt solidity highlighting disease activity. Box- and-whisker plots show corresponding quantitative AI measurements for crypt branches and crypt solidity within manual histological consensus scoring: Nancy Index remission (≤0), active disease (≥1), and inconclusive (non-agreed) cases. Visual scale showing increase disease activity in red and lower disease activity in blue. Log-transformation (log10) was applied with a +1 offset to accommodate zero values.

When the published thresholds^25^ were applied directly against Nancy consensus activity, the LP threshold had 92% sensitivity, 22% specificity and 63% accuracy, while the crypt threshold had 86% sensitivity, 57% specificity and 74% accuracy. Exploratory Nancy-derived thresholds of 70.4 cells/mm² in the LP and 24.7 cells/mm² in crypt epithelium yielded sensitivities of 65% and 72%, specificities of 87% and 93%, and accuracies of 74% and 80%, respectively.

In the strict-remission sensitivity analysis, 109 of 112 consensus RHI-remission biopsies (97%) met the additional manual component-zero criterion. At subject level, 18 of 83 participants (22%) had RHI ≤3 in every analysed biopsy and 17 of 83 (20%) also met the strict component-zero definition throughout. Active-versus-remission AUCs in the strict subset were similar to the primary analysis: 0.80 (95% CI 0.75– 0.85) for LP neutrophils, 0.86 (95% CI 0.81–0.91) for crypt neutrophils, 0.87 (95% CI 0.83–0.91) for mucin depletion, 0.89 (95% CI 0.85–0.93) for crypt roughness and 0.86 (95% CI 0.82–0.91) for reduced crypt solidity.

### Spatial distribution of inflammation in active UC

Across five 100 μm zones from the SE towards the MM, combined lymphocytes/plasma cell density was consistently elevated in histologically active UC than in histological remission: 1735 vs 1519 cells/mm^2^ in the superficial zone (MD 215, 95% CI 18–414) and 1612 vs 1089 cells/mm² in the deepest zone (MD 522, 95% CI 226–811; p<0.01 in the superficial zone and p<0.0001 in zones 200–500 μm (**Figure 4B**). This pattern was also evident across NI grades for both pathologists (**Supplementary Figure 7A/B**). Active disease showed a flatter mucosal depth gradient than histological remission (−0.047 vs -0.095 log_10_ per 100 μm; corresponding to 10% vs 20% decline respectively; slope difference 0.048 (95% CI 0.023–0.073; p<0.01), consistent with attenuated mucosal zonation.

Mucosal spatial metrics demonstrated good discrimination between active and histological remission disease (**Supplementary Figure 7C**). Mean density from SE to MM yielded an AUC of 0.69 (95% CI 0.62–0.75), and the mucosal depth gradient an AUC of 0.64 (95% CI 0.57–0.71). Combining mean density and slope increased the AUC to 0.71 (95% CI 0.64–0.77).

### Characterisation of Inconclusive UC with PAIR-IBD

Inconclusive biopsies showed mixed inflammatory profiles. Mucin depletion and surface-epithelium neutrophils were histologically remission-like, LP and crypt neutrophils were intermediate, crypt solidity and roughness were active-like, and crypt branching was greater than in either consensus group. Pairwise ROC analyses reflected this pattern: inconclusive biopsies were better separated from histological remission by crypt roughness (AUC 0.82, 95% CI 0.73–0.91), crypt branching (AUC 0.82, 95% CI 0.72–0.90), reduced crypt solidity (inverted AUC 0.82, 95% CI 0.71–0.90), crypt neutrophils (AUC 0.64, 95% CI 0.51–0.76) and crypt abscesses (AUC 0.58, 95% CI 0.48–0.68). Relative to active disease, lower mucin depletion (AUC 0.11, 95% CI 0.06–0.17), LP neutrophils (AUC 0.30, 95% CI 0.21–0.40) and surface-epithelium neutrophils (AUC 0.33, 95% CI 0.24–0.43) supported histologically remission-like positioning for those features; for mucin depletion, this corresponded to 89% (95% CI 83–94%) of inconclusive biopsies showing lower values than active biopsies using the Nancy index (87% Geboes, 81% Robarts).

## DISCUSSION

This evaluation of a new AI-system for quantification of inflammatory cell populations, crypt injury and epithelial damage, PAIR-IBD, applied to 272 UC biopsy samples from 83 subjects, revealed three features of active disease. Firstly, active UC is characterised by marked increases in LP, surface-epithelium and crypt neutrophils, together with elevated combined LP lymphocytes and plasma cells. Secondly, epithelial and architectural integrity are reduced in active disease, reflected by greater mucin depletion, increased crypt roughness, and reduced crypt solidity. Thirdly, crypt-level injury, such as branching abnormalities and abscess frequency, was prevalent in active UC than in histological remission. Most PAIR-IBD metrics demonstrated moderate-to-strong correlation with manual pathology scoring and established global indices. Together, these quantitative outputs highlight disease features that may help biologically position borderline or inconclusive cases, with mucin depletion providing the strongest support for remission-like positioning and crypt branching the clearest support for active-like positioning.

Quantifying inflammatory and architectural features across mucosal compartments distinguishes PAIR-IBD from previous AI tools, which typically assess isolated metrics or overall activity scores^25–34,40^. Such a quantitative and spatially resolved framework is well suited to clinical trial settings, where objective, reproducible assessment of histological activity and mucosal healing is required. Within this framework, neutrophil metrics and crypt abscess frequency primarily reflect active inflammatory activity, whereas chronic mononuclear inflammation and crypt branching characterise the chronic inflammatory infiltrate and chronic mucosal injury, respectively. Mucin depletion, surface-epithelium neutrophil involvement, increased crypt roughness and reduced solidity capture epithelial injury and architectural remodelling, while altered mucosal cell gradients quantify spatial organisation through mucosal depth. These domains overlap and are not intended as mutually exclusive biological states. Epithelial metaplasia, regeneration and other forms of tissue adaptation were not directly assessed.

### Inflammatory cell quantification

Neutrophil density was markedly higher in the LP, surface epithelium and crypts in active UC compared with histological remission, consistent with the established role of neutrophils as recognised markers of histological activity^41^. Our findings align with previous AI systems showing that neutrophil densities drive activity scores within the Nancy and Geboes systems^32^. A recent analysis proposed a neutrophil LP density threshold (30.4 cells/mm^2^) and crypt epithelium neutrophil density threshold (10.1 cells/mm^2^) as reliable indicators of treatment response^25^. When applied directly to our data, both thresholds were sensitive for activity, but specificity was limited, particularly for LP neutrophil. Exploratory cohort-derived thresholds provided more balanced discrimination but require external validation. Our cohort-derived threshold recovered a specificity of 86.9%, closely matching pooled field-level estimates for AI-histology models assessed against pathologists (87%, 95% CI 84–91%) from a recent meta-analysis of 12 studies (25 pooled contingency tables)^42^. This convergence suggests PAIR-IBD’s neutrophil measurements discriminate activity comparably to existing systems once appropriately calibrated; by contrast, the externally published threshold’s poor transfer to this cohort (specificity 22%) illustrates that absolute density cut-offs are unlikely to generalise directly across AI pipelines without recalibration.

Among the 112 biopsies classified as RHI remission by both pathologists, all biopsies showed residual neutrophil activity detected by PAIR-IBD. Using exploratory RHI-derived thresholds, 19 of 112 RHI-remission biopsies (17.0%) exceeded at least one threshold; these biopsies remained classified as RHI remission. These findings suggest that very subtle inflammatory features may be overlooked in routine ordinal scoring, supporting its potential role as an adjunct to manual assessment. A baseline measurement from healthy, non-IBD tissue would also be required to determine whether these residual signals reflect genuine subclinical inflammation or simply fall within the normal physiological range.

Mucosal spatial analysis revealed that combined plasma cell and lymphocytes in LP are most abundant within the superficial LP (100 μm from SE) and progressively decline toward the MM, consistent with the expected zonation of immune cells in healthy and quiescent mucosa. We found that in active UC, this organisation is markedly disrupted. The AI-based spatial profiling therefore captured spatial and architectural signatures of disease activity that conventional histological scores do not quantify^6^. Importantly, this attenuation reflects loss of normal mucosal compartmentalisation rather than cell burden alone, positioning spatial zonation as a quantitative measure of tissue organisation and inflammatory redistribution.

LP Eosinophil density did not differ significantly between active disease and remission and was numerically lower in active biopsies. Although eosinophils are less central to UC activity than neutrophils^43^, their relationship with activity is variable across cohorts^29^.

### Crypt injury and epithelial damage quantification

In active UC, PAIR-IBD quantified increases in mucin depletion, crypt roughness, crypt branching and crypt abscess formation with reductions in crypt solidity. Crypt distortion, including branching, irregularity, dilation, and variation in size and shape, is associated with higher relapse rates even among subjects in endoscopic remission^44^.

Goblet cell mucin has been shown to outperform manual scoring as a relapse predictor^31^. PAIR-IBD mucin depletion was approximately 21% higher in active disease than histological remission, consistent with the decline in goblet cell ratio decline reported in active disease^45^. Among global indices, the strongest crypt-related PAIR-IBD metrics were observed for mucin depletion and crypt neutrophils, with crypt roughness and reduced solidity also showing strong correlations with NI, SGS, and RHI.

### Associations with manual scoring

PAIR-IBD features most closely associated with overall histological activity were mucin depletion, crypt neutrophilia and crypt architectural disruption, whereas LP and surface-epithelium neutrophils showed more moderate or localised associations. In biopsies with discordant manual reads, PAIR-IBD provided exploratory biological characterisation rather than definitive adjudication. Inconclusive biopsies appeared remission-like for overt injury features, including surface-epithelial neutrophils and mucin depletion while several architectural measures remained intermediate or abnormal relative to remission. These findings suggests that AI-derived continuous metrics may support future adjudication workflows by highlighting continuous gradients of mucosal injury as decision-support measurements for pathologists. Future work exploring this utility could include confirmation against an independent pathologist or adjudicated reference before PAIR-IBD could be used to support formal case reclassification. The coexistence of remission-like or intermediate inflammatory features with persistent architectural abnormalities in inconclusive biopsies further supports mucosal healing as a biological continuum rather than a binary state.

#### Clinical implications

Histological remission is increasingly incorporated in UC drug development^46–52^ requiring expert readers, central adjudication and ordinal indices for endpoint assessment. In this context, continuous, spatially resolved AI-measurements may provide complementary quantitative information on inflammatory burden, epithelial injury and architectural abnormality. In the present study, PAIR-IBD provided exploratory biological characterisation of discordant cases, which supports a potential role as an adjunct to conventional histological scoring in future clinical trial research.

Biopsies meeting conventional remission thresholds may still show quantitatively measurable epithelial injury, architectural abnormality, or increased inflammatory cell populations in some cases. These observations should not be interpreted as redefining histological remission, but rather as evidence that AI-derived quantitative features may help characterise the spectrum of mucosal healing more granularly than ordinal scores alone. This may be particularly relevant where subtle residual abnormalities could mark incomplete healing despite formal remission. Further longitudinal studies will be required to determine whether these residual abnormalities are associated with clinical outcomes, relapse risk, or treatment response.

Future integration of AI-derived histology metrics with endoscopy, molecular readouts and biochemical biomarkers may further support multimodal disease stratification and treatment-response modelling, consistent with emerging “endo-histo-omics” frameworks for next-generation AI-enabled IBD assessment^53^. However, prospective validation in independent cohorts will be required before PAIR-IBD can be used to inform clinical decision-making, reduce adjudication burden, or improve endpoint performance in UC trials.

### Challenges and limitations

Despite significant strengths, several challenges remain. The evaluation cohort comprised 83 subjects from a single clinical network, and broader external validation across laboratories, staining protocols, scanners and disease activity spectra is required before clinical implementation. Although stain-based colour augmentation was applied during training and file formats were harmonised, explicit stain normalisation was not used. These factors may influence model performance across external settings, and future multi-site, multi-laboratory evaluation will be required before any claims of generalisability can be made. Multi-site deployments of AI histology tools have demonstrated the feasibility of this approach elsewhere, with population-diversified training across nine centres in six countries improving robustness relative to single-site development^54^; comparable external validation will be required before PAIR-IBD’s generalisability can be established. Training annotations were produced by a single expert annotator and multi-annotator validation was not undertaken. Analyses were performed at biopsy rather than subject level. Vascular lumina were not explicitly segmented, although the likely impact of intravascular cells on LP density estimates is limited in mucosal pinch biopsies where large vessels are uncommon. Nevertheless, segmentation performance remained strong across the principal tissue classes, with moderate performance for surface epithelium and muscularis mucosa, reflecting the known difficulty of segmenting these structures. Cell detection accuracy metrics reflect agreement with a single expert pathologist. Inter-rater reliability is well documented for inflammatory cell scoring but could not be quantified here, and reported accuracy may therefore be higher here than it would be against an absolute ground truth. Furthermore, the pathologist corrected a copy of the model’s own predictions rather than annotating blind, which may inflate precision through anchoring bias. Recall, driven by the pathologist actively adding missed cells, is less exposed to this bias but still reflects one individual’s judgment. Independent double-reading of a region subset would be needed to establish a firmer accuracy bound. Although ROC-based discrimination was assessed, formal classifier development and validation against independent clinical, biomarker, endoscopic and longitudinal outcomes were beyond the scope of this study. Consequently, these findings do not establish improved clinical classification, endpoint accuracy or patient-level decision-making. The current framework did not assess metaplastic epithelial phenotypes, including left-sided Paneth cell metaplasia and pyloric gland metaplasia, or explicitly model basal plasmacytosis using basal-mucosal localisation or plasma-cell clustering relative to crypt bases. Averaging plasma-cell density across the LP may therefore have limited its discriminatory performance.

Our findings demonstrate that AI-based quantification of inflammatory and architectural features provides a robust and objective means of distinguishing pathologist-defined active UC from remission. By delivering quantitative measurements across the LP, crypt epithelium and mucosal depth, PAIR-IBD complements conventional histological scoring with biologically interpretable cellular, architectural and spatial information. As digital pathology matures, such tools may support histological standardisation, adjudication and treatment-response assessment in UC clinical trials and longitudinal disease monitoring.

## Acknowledgments

We would like to thank the patients recruited into the study for their participation. We also like to thank the research nurses who collated the data at West Hertfordshire, particularly Alice Balaican. Finally, we acknowledge the IQPath facility at the UCL Queen Square Institute of Neurology for scanning the biopsy slides.

## Authors’ Contributions

Guarantor – CL. Conceptualisation – DW/AM (Equal), JL (Supporting). Writing (original draft)-DW (Lead), AM (Supporting). Data curation – AM (Lead), DW/RL/TD/CB/DW/EF/RG (Supporting). Investigation – DW (Lead), AM/TD/CB (Supporting). Formal analysis – AM/DW (Equal), TD (Supporting). Preparation – DW (Lead), AM/HTB/SL (Supporting), Visualisation – AM (Lead), DW (Supporting). Review & Editing – DW/HTB (Equal), AM/RL/TD/CD/SL/PA/RK/PW/CL/NP/MD/ACB/RF/EF/RG/JL (Supporting).

## Data availability statement

The PAIR-IBD detection system described in this study is the intellectual property of Perspectum Ltd and is the subject of a patent application (PCT/IB2025/062039). Summary data are included in the manuscript or uploaded as supplemental information. Additional data underlying this study are proprietary and not publicly available, but may be considered for sharing on a case-by-case basis for legitimate research or commercial collaboration, subject to a data sharing agreement and applicable governance and ethical approvals. Requests should be directed to the corresponding author.

## Ethics approval statement

All parent studies were conducted in accordance with the ethical principles of the Declaration of Helsinki 2013 and the Good Clinical Practice guidelines and all studies approved by the London-Queen Square Research Ethics Committee (REC: 20/LO/0349).

## Patient consent statement

Informed consent and assent (where required) was provided by all subjects and their caregivers (where required), respectively prior to their participation in the study.

## Permission to reproduce material from other sources

Anonymised individual participant data can be shared upon request or as required by law and/or regulation with qualified external researchers. Approval of such requests is at the discretion of the study sponsors and is dependent on the nature of the request, the merit of the research proposed, the availability of the data, and the intended use of the data.

## Clinical trial registration

The WHITS study was registered as NCT05000242.

