## Supplementary Information for "Spatial AI quantification of inflammatory and architectural features differentiates pathologist-defined active ulcerative colitis from histological remission"

*Study Design and Populations*

Recruitment exclusion criteria included female participants who are, or suspect that they are, pregnant at the time of clinical assessment and any other cause, including significant disease or disorder which may either put the participant at risk because of participation in the study, or influence the participant’s ability to participate in the study. Participants were enrolled during routine clinical visits across two sites within the West Hertfordshire Hospitals NHS Trust (Hemel Hempstead and Watford endoscopy units).

*Blood tests, stool and biopsy sample collection*

Samples were restricted to the large intestine and any other biopsies collected were not analyzed (e.g. small intestine, tubular adenoma, etc.). Any areas that did not contain bowel tissue or were not stained with Hematoxylin & Eosin were removed. C-Reactive Protein (CRP; normal <5 mg/L), F-Calprotectin (normal <50 μg/g), albumin (normal <40 g/L) and haemoglobin (normal range: 165-180 g/L for men and 120-165 g/L for women) were collected with standard methods in accredited clinical laboratories. Treatment for UC was considered as ongoing with either topical, biologic/small molecule or only steroids.

*Imaging Protocol*

Whole Slide Imaging H&E and IBD biopsy slides were digitized using two scanners. Slides captured as SVS scans were acquired using a Pannoramic® 480 3DHistech scanner at 40x (243 nm/pixel; 104703 DPI). Slides captured as NDPI scans were acquired using a Hamamatsu NanoZoomer® S360 Digital slide scanner C13220-04 at 40x (230 nm/pixel; 110208 DPI). SVS and NDPI files were converted to TIFF to ensure 0.5mpp resolution in the processed image. Stain-based colour augmentation was applied during training to improve robustness to staining variability across slides and scanners.

Each whole-slide image (WSI) contained three serial sections with multiple pinch biopsies per slide. AI-based quantification of inflammatory cell, crypt injury and epithelial damage was performed on the biopsy section with the highest lymphocyte density as determined by PAIR-IBD (**Figure 1**). Minimum area of LP was determined to be 1mm^2^ and WSIs under this threshold were removed from analysis. Images that were of poor quality (blurry, not enough biopsy) were manually removed prior to AI analysis.

*Pathologist scoring*

At baseline, biopsies per subject ranged from 1-7 and were scored independently and individually, rather than aggregated at the subject level. Two pathologists scored 272 UC biopsies using a universal scoring template (**Supplementary Table 1**). Established reading plans and alignment meetings were used to increase agreement between the two pathologists. The universal scoring template enabled the conversion into 3 scoring indices (Simplified Geboes Score, Robarts Histopathological Score and Nancy Index).

*Biopsy Quality Control*

PAIR-IBD is trained to segment high quality tissue foreground areas using a large corpus of tissue types and common scanning artefacts and the system rejects areas of text, black ink marks and other artefacts from downstream analyzes. PAIR-IBD links tissue features to their parent section of tissue allowing spatially localized quantification of IBD metrics. To ensure sufficient tissue was analyzed per slide we chose a minimum area criterion of 1mm^2^ of lamina propria per analyzed section.

*Algorithm pipeline*

PAIR-IBD was developed for H&E image analysis, enabling the spatial quantification of both non-pathological and pathological features relevant to IBD activity (e.g. neutrophils, crypts, muscularis mucosa). PAIR-IBD consists of an ensemble of U-net++ convolutional neural networks trained to detect key primary histological features and derive secondary features. Target features included colorectal crypts, LP, surface epithelium, muscularis mucosa, mucin, abscesses, and blood, as well as cell types relevant to IBD pathology (neutrophils, eosinophils, lymphocytes, plasma cells, epithelial cells and fibroblasts). The crypt segmentation model delineated the external epithelial boundary, internal lumen, and mucin content. Spatial algorithms were implemented in Python using the Geopandas library^1^. Training, validation and held-out testing of segmentation models was performed on slide-level separated datasets drawn from the WHITS IBD study, reflecting the practical constraint that this was the principal cohort available during model development. Each case contributed multiple slides from distinct biological locations representing a broad spectrum of disease activity, resulting in substantial within-case heterogeneity; the training and testing datasets were therefore designed as biologically representative subsets of this broader cohort, capturing the spectrum of histological variation available. Because complete independence at the cohort level was not feasible, we focused on preventing direct sample reuse in model evaluation by enforcing exact-slide separation and auditing overlap at both slide and case resolution. There was no slide-level overlap between the training and held-out testing sets, although a subset of cases contributed slides to both sets: of 55 WHITS UC cases considered for segmentation model development, 7 contributed slides only to the training set, 9 contributed slides to both the training and testing sets, 10 contributed slides only to the testing set, and 29 contributed no slides to either set. Most WHITS slides used for downstream biological analyses were not used in segmentation model development. Additional labelled training data for crypt and surface epithelium segmentation were obtained from the CHOC and NTNU public IBD cohorts^2,3^.

Cell detection, including inflammatory cell types, employed HoVerNext pretrained weights for colorectal cell types^4^ while additional models were trained with expert-labelled annotations (62,000 image-annotation pairs; 80% training, 20% validation) from in-house (WHITS study) and publicly available data^2,3^.

To improve detection performance on rare cell types (neutrophils and eosinophils), we used an ensemble combining independent inference from HoVerNext and an in-house UNet++ model trained specifically on these cell types, using a proprietary dataset of >6,500 neutrophil and >5,000 eosinophil annotations. Predicted cell centroids from the two models were matched using the Hungarian algorithm: unmatched predictions were retained as identified by the originating model; matched predictions with concordant identity were combined into a single prediction; and matched predictions with discordant identity were resolved via a confidence-weighted voting scheme based on the in-house model.

Training annotations were performed in QuPath by an experienced annotator. Annotations were exported as pixel-level masks and used to supervise convolutional neural network training.

*Assessment of inflammatory cell identification accuracy*

To assess the performance of our inflammatory cell identification model, an expert GI pathologist reviewed the model's cell-level point predictions across 17 tissue regions from 17 slides from the study cohort. Assessment regions were selected to reflect a range of inflammatory cell abundances.

The pathologist's review was scoped to four inflammatory cell classes: eosinophils, neutrophils, plasma cells, and lymphocytes, reflecting the inflammatory metrics reported in this paper. For these classes, we created a system that allowed the reviewer to confirm correct detections in place, alter the position, delete spurious (false-positive) predictions, correct misclassified points by relabeling them, and add new point annotations for any inflammatory cells the model had missed. Predictions for non-inflammatory cell classes present in PAIR-IBD’s output (e.g. epithelial cells, connective-tissue cells, goblet cells) were not reviewed and are excluded from the performance analysis below.

Model predictions were compared against the reviewed reference points using a distance-constrained bipartite matching procedure. For each region, predicted and reference points were matched using the Hungarian algorithm, restricted to candidate pairs within a maximum Euclidean distance of 15 pixels (~7 µm), with candidates identified efficiently via a k-d tree. Matching was performed twice per region: once with class-aware constraints (a match required identical predicted and reference class labels), used to compute per-class precision, recall, and F1 score; and once class-agnostically (matching by location alone), used to separately quantify detection performance independent of classification, and to construct a confusion matrix over the four reviewed classes that explicitly accounts for missed detections (unmatched reference points) and spurious detections (unmatched predictions).

For each class, we report micro-averaged precision, recall, and F1 score, pooling true positives, false positives, and false negatives across all reviewed regions. Ninety-five percent confidence intervals were obtained by bootstrap resampling of regions (1,000 resamples), treating the region as the unit of statistical independence.

*Assessment of tissue feature segmentation accuracy*

We evaluated segmentation performance across six tissue classes, tissue foreground, lamina propria, crypts, muscularis mucosa, surface epithelium, and blood, in 26 WSI at 0.5 µm/px. Because most tissue classes are represented as amorphous regions rather than countable instances, we adopted a tile-based (area-level) assessment as the primary evaluation strategy. A regular 512 × 512 pixel grid was overlaid on each slide and, for every tile containing ground-truth annotations of a given class, we computed area-based precision, recall, IoU, and Dice from the intersection of dissolved ground-truth and prediction polygons with each tile. Tiles without ground-truth annotations for a class were excluded from evaluation of that class to avoid false-positive bias in regions where annotators had not labelled all instances. This yielded between 2,085 and 11,222 evaluated tiles per class (39,831 in total), providing spatially resolved, continuous metrics that are robust to differences in polygon fragmentation between ground truth and model output. We report micro-averaged metrics (area-weighted aggregation across all tiles) with 95% bootstrap confidence intervals obtained by resampling over slides (1,000 iterations) to preserve within-slide spatial correlation.

For crypts, the only class comprising discrete, countable instances (96–1,109 per slide), we additionally performed instance-level evaluation. MultiPolygon features were exploded into individual polygon instances. To ensure consistency with the tile-based evaluation, predicted instances were first spatially filtered to retain only those intersecting the union of ground-truth annotations for that slide-region, excluding predictions in regions that had not been annotated. Predicted instances were then matched one-to-one to ground-truth instances using greedy intersection-over-union (IoU) matching at a threshold of 0.5. From the matched pairs we computed Panoptic Quality (PQ), defined as the product of segmentation quality (mean matched IoU) and recognition quality (F1 of the matching).

Tile-based assessments are shown in Supplementary Table 3 and crypt instance-level segmentation in Supplementary Table 4.

*Crypt Segmentation Features*

Upon segmentation of distinct tissue domains (Lamina propria, muscularis mucosa, submucosal muscle layers, crypts and surface epithelium), we applied custom spatial algorithms to associate pertinent disease features with each domain of the tissue. Distinct regions of each tissue domain were treated separately, i.e. metrics were computed per region of lamina propria, and specific for every crypt. Crypts were associated with the surrounding piece of lamina propria by tracing all tissue subdomains back to their parent piece of tissue, also via custom spatial algorithms.

*Crypt solidity* Measure of architectural integrity; deviation of crypt structure from typical convex shape. Ratio of the crypt area to the crypt convex hull area. The convex hull is the smallest convex shape that completely encloses an object.

$$Crypt solidity=\frac{{area}_{crypt}}{{area}_{crypt convex hull}}$$

*Crypt roughness* Measure of surface irregularity along crypt epithelium.

$$Crypt roughness=\frac{{perimeter}_{crypt}}{{perimeter}_{crypt convex hull}}$$

These are assessed through analysis of the coordinates of the perimeter of the crypt describing a mathematical closed curve. This is quantified by elliptic Fourier and principal component analysis (PCA) of the closed curve describing the perimeter of the crypt. The morphological descriptors of the geometry of the crypts are then produced (e.g. area, solidity, ellipticity, roughness). This also includes the segmentation of the interior of colorectal crypts, comprising the crypt lumen and mucin pockets.

*Mucin depletion* Measure of loss of mucin content within each segmented crypt across the analyzed tissue section. Fractional mucin content was first approximated as the ratio of the internal crypt area (mucin pockets plus lumen), segmented by a model devoted to this task, to the whole crypt area determined by our crypt segmentation model. Mucin depletion was determined as 1 minus the mucin fraction. The mean of all crypts was calculated across the analyzed tissue section.

$$Mucin depletion=1-\left( \frac{{area}_{crypt interior}}{{area}_{crypt}} \right)$$

The difference between crypt geometries and crypt interior geometries is measured, for example by measuring the average distance between a radial point on the crypt interior and its equivalent point on the crypt geometry. The crypt architecture and properties of cells situated within the crypt are quantified and classified.

*Crypt abscesses* Fraction of crypts that contain a segmented abscess across the analyzed tissue section.

$$Crypt abscesses=\frac{{number}_{crypt abscess}}{{number}_{crypt}}$$

*Crypt Branching* Enumeration of the number of branches in a crypt’s morphological skeleton, measured using the Python Skan skeleton analysis package^5^. Higher branching scores indicate a deviation from normal round or test-tube like crypt structure into complex morphologies that exhibit bifurcation and abnormal branching. The AI system measures crypt structural abnormality is quantified, for example, the overall geometry of a crypt is transformed into its morphological skeleton and delineated by their external epithelial cell layer. The morphological skeleton is used to create an undirected graph representation where each segment of the skeleton is an edge in the graph. Branching crypts are detected through analysis of the graph representation (e.g. through node degree, connectedness, centrality or combinations of descriptors).

$$Crypt branches=mean n{umber}_{branches}$$

*Statistical modelling of zonal immune cell gradients* The decline in lymphocyte and plasma cell density across the tissue depth was quantified as a gradient, defined as the slope of log-transformed cell density across five anatomical zones extending from the surface epithelium to the MM. For each zone, cell density (y) was transformed as log_10_($\mathbb{y}$ + 1) to stabilise variance and accommodate zero counts. Tissue depth (“distance”) was treated as a continuous variable (in micrometres). To evaluate whether this gradient differed by histological disease activity, a linear mixed-effects model was fitted with fixed effects for distance, disease status (Active, Remission, Inconclusive), and their interaction, and a random intercept for slide to account for clustering of zones within images:

$$\log10\left( \mathbb{y}+1 \right)=\beta_{0}+\beta_{1}distance+\beta_{2}Disease Status+\beta_{3}\left( distance x Disease Status \right)+ \upsilon_{slide}+ \epsilon$$

Distance captured the change in cell density with increasing tissue depth, disease status represented differences in baseline density between activity groups, and the interaction term quantified whether the rate of decline (gradient) varied by disease state. In this model, ‘β_0_’ is the intercept (baseline log-density in the reference disease group), ‘β_1_’ is the slope describing how density changes with depth in the reference group, ‘β_2_’ represents differences in baseline density between disease-status groups, and ‘β_3_’ represents differences in slope (gradient) between disease-status groups. The slide-level random effect ($\upsilon$_slide_) absorbed between-slide variation in baseline density, ‘ε’ represented residual within-slide variability. The significance of fixed effects was evaluated using Type III ANOVA with Satterthwaite-approximated degrees of freedom.

Normally distributed continuous variables are presented as mean ± standard deviation (SD), and categorical variables as counts and percentages. Inter-rater reliability between pathologists was assessed using pairwise overall percentage agreement (OPA) and weighted Cohen’s kappa (κ), with ICC used for RHI where appropriate.

Correlations between measurements were investigated using Spearman’s rank correlations (one continuous variable, the other categorical) or Pearson’s correlation (both continuous variables) with correlations greater than 0.60 considered strong^6^. Correlations of <0.3 were considered weak and 0.3-0.6 moderate. F1 scores greater than 0.8 considered good^7^.

All statistical analyses were performed in RStudio version 4.5.1.

Supplementary Figure 1: Study design and populations

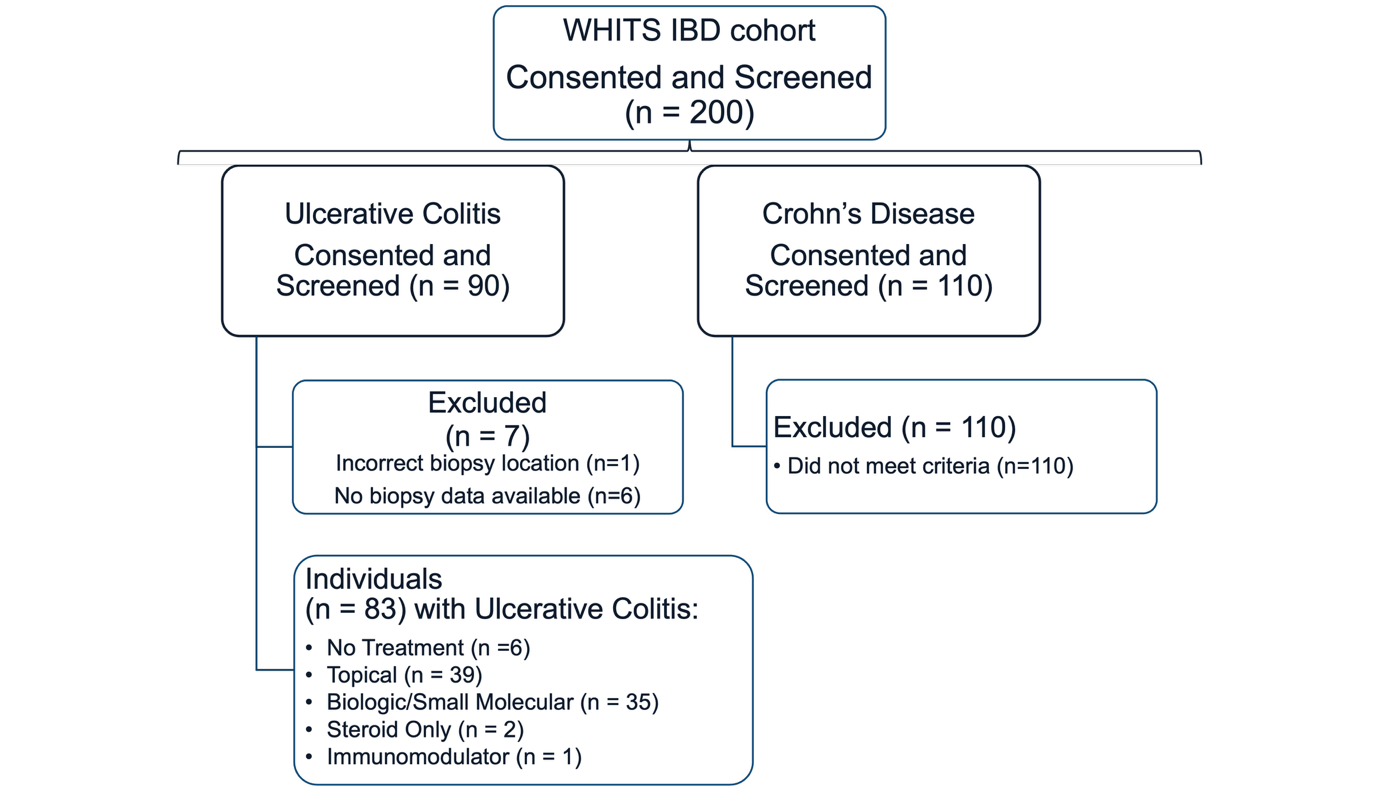

Supplementary Table 1: Universal Histological Scoring Template

| **Feature** | **Score** | **Extent** | **Scoring conversion** | **Simplified Geboes Score** | **Nancy Index** | | **Robarts Histopathology Index** |
| --- | --- | --- | --- | --- | --- | --- | --- |
| **Architectural Changes** | 0 | None | Converts Into | 0_0 |  | | |
|  | 1 | Mild |  | 0_1 |  |  |  |
|  | 2 | Moderate |  | 0_2 |  |  |  |
|  | 3 | Marked |  | 0_2 |  |  |  |
| **Serrated Architecture** | 0 | None | Does not convert into a histological score (Presence indicates higher risk of Colorectal Cancer) | | | | |
|  | 1 | <5% |  |  |  |  |  |
|  | 2 | 5-50% |  |  |  |  |  |
|  | 3 | >50% |  |  |  |  |  |
| **Mucin Depletion** | 0 | None | Converts into full Geboes Score (ECCO consensus definition and Riley Score) | | | | |
|  | 1 | Mild |  |  |  |  |  |
|  | 2 | Moderate |  |  |  |  |  |
|  | 3 | Severe |  |  |  |  |  |
| **Chronic Nuclear Infiltrate** | 0 | None | Converts Into |  | 0 | 0 | |
|  | 1 | Mild |  |  | 1 | 3 | |
|  | 2 | Moderate |  |  | 1 | 6 | |
|  | 3 | Severe |  |  | 1 | 9 | |
| **Basal Plasma Cell Increase** | 0 | None | Converts Into | 1_0 | 0 |  | |
|  | 1 | Mild |  | 1_1 | 1 |  |  |
|  | 2 | Moderate |  | 1_1 | 1 |  |  |
|  | 3 | Severe |  | 1_2 | 1 |  |  |
| **Increase in Eosinophils in LP** | 0 | None | Converts Into | 2A_0 |  | | |
|  | 1 | Mild |  | 2A_1 |  |  |  |
|  | 2 | Moderate |  | 2A_1 |  |  |  |
|  | 3 | Severe |  | 2A_2 |  |  |  |
| **Increase in Neutrophils in LP** | 0 | None | Converts Into | 2B_0 |  | 0 | |
|  | 1 | Mild |  | 2B_1 |  | 3 | |
|  | 2 | Moderate |  | 2B_1 |  | 6 | |
|  | 3 | Severe |  | 2B_2 |  | 9 | |
| **Increase in Neutrophils in Crypt Epithelium** | 0 | None | Converts Into | 3_0 | - |  | |
|  | 1 | <5% |  | 3_1 | 2 |  |  |
|  | 2 | 5-50% |  | 3_1 | 3 |  |  |
|  | 3 | >50% |  | 3_2 | 3 |  |  |
| **Crypt Abscess** | 0 | None | Converts Into |  | - |  | |
|  | 1 | <5% |  |  | 2 |  |  |
|  | 2 | 5-50% |  |  | 3 |  |  |
|  | 3 | >50% |  |  | 3 |  |  |
| **Overall Crypt Inflammation** | 0 | None | Converts Into |  |  | 0 | |
|  | 1 | <5% |  |  |  | 3 | |
|  | 2 | 5-50% |  |  |  | 6 | |
|  | 3 | >50% |  |  |  | 9 | |
| **Crypt Injury** | 0 | None | Does not convert into a histological score | | | | |
|  | 1 | Marked Attenuation |  |  |  |  |  |
|  | 2 | Probable crypt destruction |  |  |  |  |  |
|  | 3 | Definite/Unequivocal crypt destruction |  |  |  |  |  |
| **Surface Epithelial Injury** | 0 | None | Converts Into | 4_0 | - | 0 | |
|  | 1 | Marked Attenuation |  | 4_1 | - | 3 | |
|  | 2 | Probably Erosion |  | 4_2 | - | 3 | |
|  | 3 | Definite/Unequivocal erosion |  | 4_3 | 4 | 6 | |
|  | 4 | Ulcer/Granulation Tissue |  | 4_4 | 4 | 9 | |

Supplementary Table 2: Individual histological component scores by pathologist reader.

*Values are n(%) unless otherwise stated.

|  | **All (83 subjects; 272 biopsies)** |
| --- | --- |
| **Increase in Neutrophils in the LP*** | |
| Score 0 – (P1-P2) | 176 (65%) – 227 (83%) |
| Score 1 – (P1-P2) | 86 (32%) – 37 (14%) |
| Score 2 – (P1-P2) | 9 (3%) – 7 (3%) |
| Score 3 – (P1-P2) | 1 (<1%) – 1 (<1%) |
| **Increase in Neutrophils in Crypt Epithelium*** | |
| Score 0 – (P1-P2) | 139 (51%) – 131 (48%) |
| Score 1 – (P1-P2) | 76 (28%) – 88 (32%) |
| Score 2 – (P1-P2) | 52 (19%) – 33 (12%) |
| Score 3 – (P1-P2) | 5 (2%) – 20 (7%) |
| **Increase in Basal Plasma Cells*** | |
| Score 0 – (P1-P2) | 158 (58%) – 119 (44%) |
| Score 1 – (P1-P2) | 68 (25%) – 74 (27%) |
| Score 2 – (P1-P2) | 30 (11%) – 32 (12%) |
| Score 3 – (P1-P2) | 16 (6%) – 47 (17%) |
| **Increase in Eosinophils in the LP*** | |
| Score 0 – (P1-P2) | 196 (72%) – 209 (77%) |
| Score 1 – (P1-P2) | 72 (26%) – 53 (19%) |
| Score 2 – (P1-P2) | 4 (1%) – 8 (3%) |
| Score 3 – (P1-P2) | 0 – 2 (<1%) |
| **Chronic Mononuclear Infiltrate*** | |
| Score 0 – (P1-P2) | 101 (37%) – 120 (44%) |
| Score 1 – (P1-P2) | 90 (33%) – 44 (16%) |
| Score 2 – (P1-P2) | 64 (24%) – 57 (21%) |
| Score 3 – (P1-P2) | 17 (6%) – 51 (19%) |
| **Architectural Changes*** | |
| Score 0 – (P1-P2) | 83 (31%) – 122 (45%) |
| Score 1 – (P1-P2) | 127 (47%) – 64 (24%) |
| Score 2 – (P1-P2) | 47 (17%) – 37 (14%) |
| Score 3 – (P1-P2) | 15 (6%) – 49 (18%) |
| **Serrated Architecture*** | |
| Score 0 – (P1-P2) | 244 (90%) – 222 (82%) |
| Score 1 – (P1-P2) | 25 (9%) – 37 (14%) |
| Score 2 – (P1-P2) | 3 (1%) – 8 (3%) |
| Score 3 – (P1-P2) | 0 (0%) – 5 (2%) |
| **Crypt Injury*** | |
| Score 0 – (P1-P2) | 215 (79%) – 229 (84%) |
| Score 1 – (P1-P2) | 19 (7%) – 12 (4%) |
| Score 2 – (P1-P2) | 26 (10%) – 12 (4%) |
| Score 3 – (P1-P2) | 12 (4%) – 19 (7%) |
| **Overall Crypt Inflammation*** | |
| Score 0 – (P1-P2) | 137 (50%) – 133 (49%) |
| Score 1 – (P1-P2) | 63 (23%) – 74 (27%) |
| Score 2 – (P1-P2) | 60 (22%) – 44 (16%) |
| Score 3 – (P1-P2) | 12 (4%) – 21 (8%) |
| **Mucin Depletion*** | |
| Score 0 – (P1-P2) | 116 (43%) – 169 (62%) |
| Score 1 – (P1-P2) | 82 (30%) – 52 (19%) |
| Score 2 – (P1-P2) | 62 (23%) – 39 (14%) |
| Score 3 – (P1-P2) | 12 (4%) – 12 (4%) |
| **Crypt Abscess*** | |
| Score 0 – (P1-P2) | 201 (74%) – 200 (74%) |
| Score 1 – (P1-P2) | 44 (16%) – 46 (17%) |
| Score 2 – (P1-P2) | 25 (9%) – 24 (9%) |
| Score 3 – (P1-P2) | 2 (1%) – 2 (1%) |
| **Epithelial Injury*** | |
| Score 0 – (P1-P2) | 200 (74%) – 222 (82%) |
| Score 1 – (P1-P2) | 37 (14%) – 23 (8%) |
| Score 2 – (P1-P2) | 8 (3%) – 7 (3%) |
| Score 3 – (P1-P2) | 17 (6%) – 16 (6%) |
| Score 4 – (P1-P2) | 10 (4%) – 4 (1%) |

Supplementary Table 3: Tile-based segmentation performance by tissue class

| **Tissue class** | **n tiles** | **Dice (95% CI)** | **IoU (95% CI)** |
| --- | --- | --- | --- |
| **Tissue foreground** | 11,222 | 0.990 (0.989–0.991) | 0.980 (0.978–0.983) |
| **Lamina propria** | 11,190 | 0.921 (0.907–0.933) | 0.855 (0.831–0.876) |
| **Crypts** | 8,482 | 0.919 (0.902–0.932) | 0.850 (0.822–0.873) |
| **Blood** | 2,085 | 0.680 (0.636-0.707) | 0.772 (0.742-0.800) |
| **Muscularis mucosa** | 2,118 | 0.809 (0.769-0.855) | 0.679 (0.624-0.746) |
| **Surface epithelium** | 4,734 | 0.719 (0.675–0.753) | 0.561(0.509–0.604) |

Supplementary Table 4: Crypt instance-level segmentation performance

| **Metric** | **Value** |
| --- | --- |
| **Panoptic Quality (PQ)** | 0.703 |
| **Precision** | 0.848 |
| **Recall** | 0.763 |
| **F1 score** | 0.802 |
| **Mean matched IoU** | 0.874 |

Supplementary Table 5: Cell-level detection performance versus pathologist review.

*Additional cells of each type identified by the pathologist reviewer but not detected by the model, across 17 reviewed regions.

| **Cell Type** | **Precision** | **Recall** | **F1 (95% CI)** | **Additional cells identified by pathologist*** |
| --- | --- | --- | --- | --- |
| **All classes  (class-agnostic)** | 0.998 | 0.934 | 0.965 (0.945–0.980) | 169 |
| **Lymphocytes** | 1.000 | 0.992 | 0.996 (0.990–0.999) | 16 |
| **Eosinophils** | ≥0.989 | 0.779 | (0.818–0.930) | 49 |
| **Neutrophils** | ≥0.989 | 0.776 | (0.816–0.921) | 46 |
| **Plasma cells** | ≥0.989 | 0.756 | (0.726–0.924) | 58 |

Supplementary Table 6: AI metric overview with pooled metrics from two pathologists – Nancy Index

| **Metric** | **All# (n=272)** | **Remission#** | **Active#** | **Inconclusive#** | **Remission – Active  (p value)** | **Active – Inconclusive**  **(p value)** | **Remission – Inconclusive**  **(p value)** |
| --- | --- | --- | --- | --- | --- | --- | --- |
| **Neutrophils (LP)** | 78.37 ± 57.88 | 49.54 ± 28.78 | 101.67 ± 65.84 | 61.13 ± 36.77 | p < 0.0001 | p < 0.01 | p = 0.55 |
| **Eosinophils** | 153.47 ± 124.27 | 167.18 ± 130.20 | 141.21 ± 113.49 | 167.53 ± 150.70 | p = 0.26 | p = 1 | p = 1 |
| **Lymphocytes (LP)** | 2277.91 ± 759.97 | 2021.59 ± 642.03 | 2490.19 ± 818.94 | 2098.84 ± 474.12 | p < 0.0001 | p < 0.05 | p = 1 |
| **Plasma Cells (LP)** | 137.01 ± 145.72 | 137.52 ± 165.52 | 134.27 ± 136.90 | 148.89 ± 117.22 | p = 1 | p = 0.52 | p = 0.72 |
| **Crypt Neutrophils** | 36.92 ± 45.07 | 10.92 ± 8.49 | 57.90 ± 52.24 | 21.54 ± 23.88 | p < 0.0001 | p < 0.0001 | p = 0.072 |
| **Mucin Depletion** | 0.78 ± 0.11 | 0.70 ± 0.08 | 0.85 ± 0.09 | 0.70 ± 0.07 | p < 0.0001 | p < 0.0001 | p = 1 |
| **Crypt Abscess** | 0.03 ± 0.06 | 0.01 ± 0.03 | 0.05 ± 0.07 | 0.02 ± 0.06 | p < 0.0001 | p < 0.05 | p = 0.21 |
| **Crypt Branches** | 1.43 ± 0.81 | 1.04 ± 0.50 | 1.56 ± 0.75 | 2.10 ± 1.21 | p < 0.0001 | p = 0.08 | p < 0.0001 |
| **Crypt Solidity** | 0.88 ± 0.06 | 0.92 ± 0.03 | 0.85 ± 0.06 | 0.86 ± 0.06 | p < 0.0001 | p = 0.93 | p < 0.0001 |
| **Crypt Roughness** | 1.14 ± 0.08 | 1.08 ± 0.04 | 1.18 ± 0.08 | 1.16 ± 0.10 | p < 0.0001 | p = 0.79 | p < 0.0001 |

Supplementary Table 7: AI metric overview with pooled metrics from two pathologists – Robarts Histopathological Index

| **Metric** | **All# (n=272)** | **Remission#** | **Active#** | **Inconclusive#** | **Remission – Active  (p value)** | **Active – Inconclusive**  **(p value)** | **Remission – Inconclusive**  **(p value)** |
| --- | --- | --- | --- | --- | --- | --- | --- |
| **Neutrophils (LP)** | 78.37 ± 57.88 | 48.90 ± 27.71 | 103.54 ± 65.69 | 65.37 ± 40.03 | p < 0.0001 | p < 0.05 | p = 0.38 |
| **Eosinophils** | 153.47 ± 124.27 | 162.52 ± 128.55 | 147.22 ± 122.60 | 146.46 ± 113.28 | p = 0.7 | p = 1 | p = 1 |
| **Lymphocytes (LP)** | 2277.91 ± 759.97 | 2030.23 ± 614.71 | 2503.84 ± 811.70 | 2061.26 ± 640.12 | p < 0.0001 | p = 0.059 | p = 1 |
| **Plasma Cells (LP)** | 137.01 ± 145.72 | 142.87 ± 161.85 | 136.45 ± 139.23 | 106.59 ± 78.72 | p = 1 | p = 1 | p = 1 |
| **Crypt Neutrophils** | 36.92 ± 45.07 | 10.99 ± 8.52 | 58.71 ± 52.40 | 28.07 ± 28.13 | p < 0.0001 | p < 0.05 | p < 0.01 |
| **Mucin Depletion** | 0.78 ± 0.11 | 0.70 ± 0.08 | 0.84 ± 0.09 | 0.73 ± 0.08 | p < 0.0001 | p < 0.0001 | p = 0.61 |
| **Crypt Abscess** | 0.03 ± 0.06 | 0.01 ± 0.03 | 0.05 ± 0.07 | 0.02 ± 0.04 | p < 0.0001 | p = 0.089 | p = 0.1 |
| **Crypt Branches** | 1.43 ± 0.81 | 1.09 ± 0.52 | 1.65 ± 0.88 | 1.77 ± 0.94 | p < 0.0001 | p = 1 | p < 0.01 |
| **Crypt Solidity** | 0.88 ± 0.06 | 0.92 ± 0.03 | 0.84 ± 0.06 | 0.87 ± 0.06 | p < 0.0001 | p = 0.61 | p < 0.01 |
| **Crypt Roughness** | 1.14 ± 0.08 | 1.08 ± 0.04 | 1.18 ± 0.08 | 1.15 ± 0.08 | p < 0.0001 | p = 0.36 | p < 0.001 |

Supplementary Table 8: AI metric overview with pooled metrics from two pathologists – Simplified Geboes Score

| **Metric** | **All# (n=272)** | **Remission#** | **Active#** | **Inconclusive#** | **Remission – Active  (p value)** | **Active – Inconclusive**  **(p value)** | **Remission – Inconclusive**  **(p value)** |
| --- | --- | --- | --- | --- | --- | --- | --- |
| **Neutrophils (LP)** | 78.37 ± 57.88 | 50.20 ± 31.48 | 103.52 ± 66.34 | 75.09 ± 40.80 | p < 0.0001 | p = 0.18 | p < 0.01 |
| **Eosinophils** | 153.47 ± 124.27 | 166.88 ± 129.00 | 133.90 ± 103.73 | 198.99 ± 182.34 | p = 0.11 | p = 0.71 | p = 1 |
| **Lymphocytes (LP)** | 2277.91 ± 759.97 | 2029.58 ± 618.85 | 2494.09 ± 816.19 | 2280.25 ± 712.60 | p < 0.0001 | p = 0.61 | p = 0.42 |
| **Plasma Cells (LP)** | 137.01 ± 145.72 | 140.83 ± 161.47 | 127.05 ± 130.66 | 175.37 ± 143.62 | p = 0.99 | p = 0.087 | p = 0.29 |
| **Crypt Neutrophils** | 36.92 ± 45.07 | 11.14 ± 8.68 | 60.58 ± 53.12 | 30.15 ± 27.52 | p < 0.0001 | p < 0.05 | p < 0.01 |
| **Mucin Depletion** | 0.78 ± 0.11 | 0.70 ± 0.08 | 0.86 ± 0.08 | 0.72 ± 0.09 | p < 0.0001 | p < 0.0001 | p = 1 |
| **Crypt Abscess** | 0.03 ± 0.06 | 0.01 ± 0.03 | 0.05 ± 0.07 | 0.04 ± 0.08 | p < 0.0001 | p = 0.19 | p < 0.05 |
| **Crypt Branches** | 1.43 ± 0.81 | 1.15 ± 0.60 | 1.53 ± 0.74 | 2.25 ± 1.30 | p < 0.0001 | p < 0.05 | p < 0.0001 |
| **Crypt Solidity** | 0.88 ± 0.06 | 0.91 ± 0.03 | 0.85 ± 0.06 | 0.85 ± 0.07 | p < 0.0001 | p = 1 | p < 0.0001 |
| **Crypt Roughness** | 1.14 ± 0.08 | 1.08 ± 0.04 | 1.18 ± 0.08 | 1.20 ± 0.10 | p < 0.0001 | p = 1 | p < 0.0001 |

Supplementary Figure 2 – AI derived metrics compared in samples with different grades from manual reads (Universal Scoring Template). Neutrophil density in the LP (A), crypt neutrophil density (B) and lymphocyte density in the LP are compared to each individual pathologist’s grading in UC (n = 272).

**A)**

**C)**

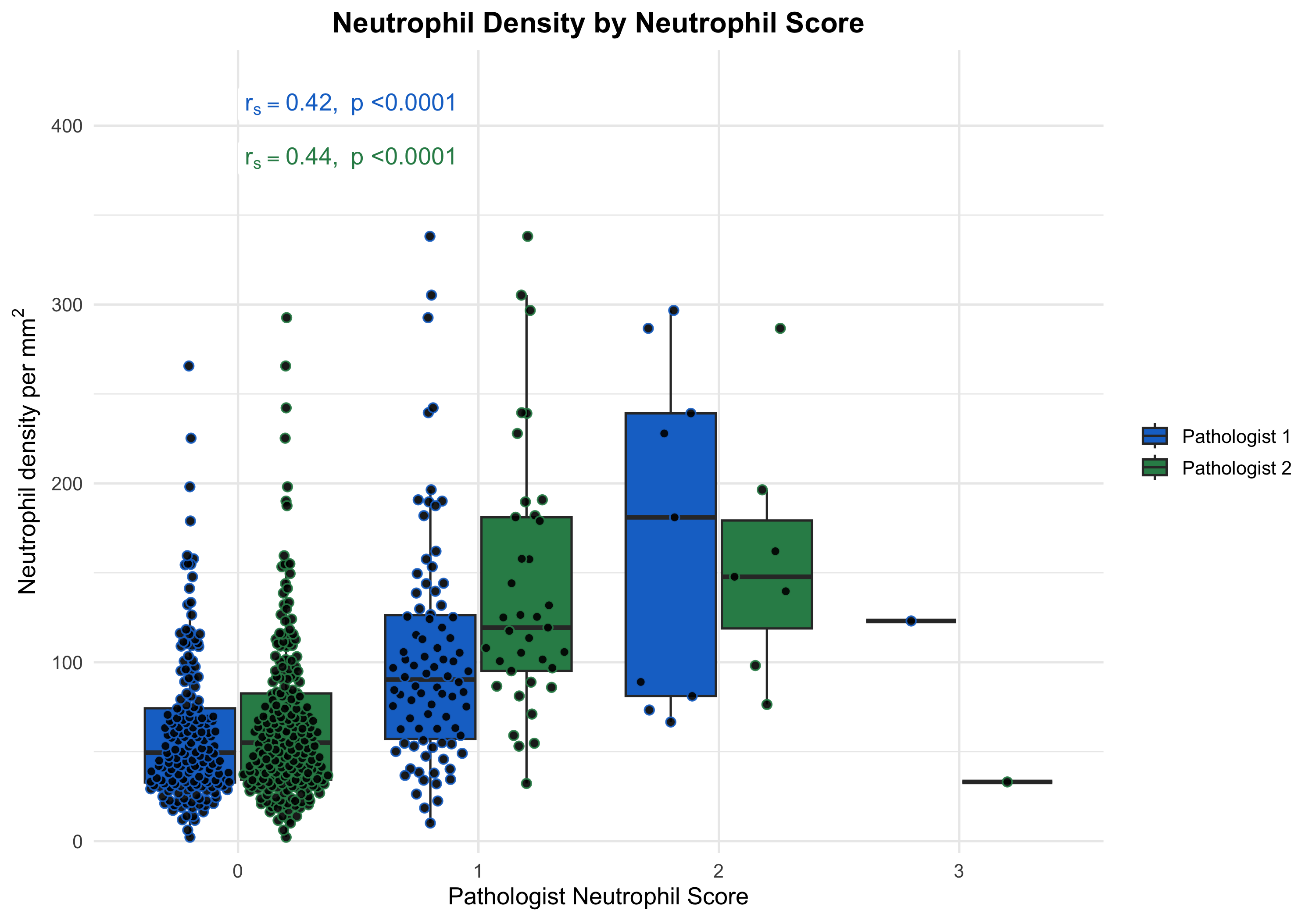

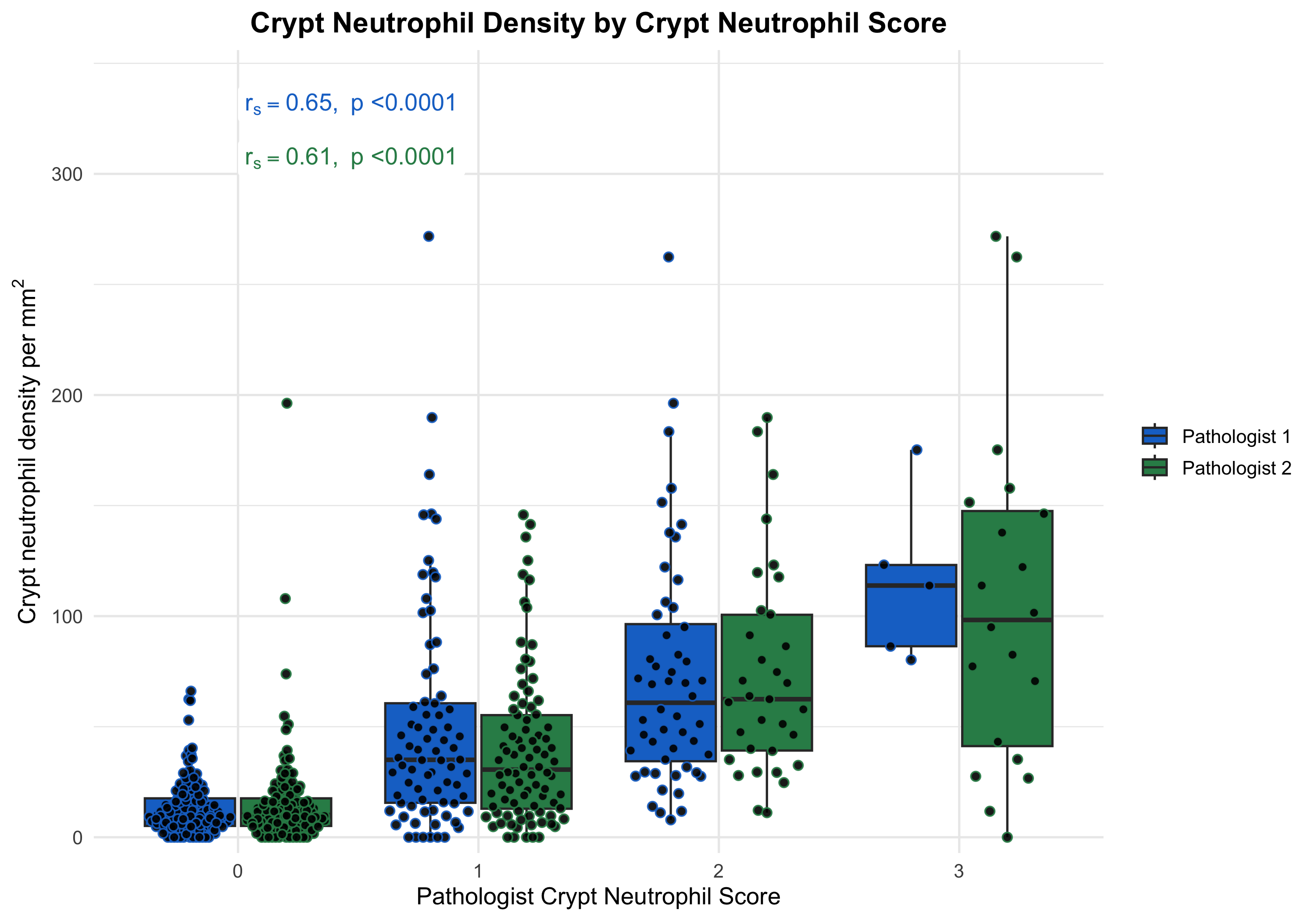

**B)**

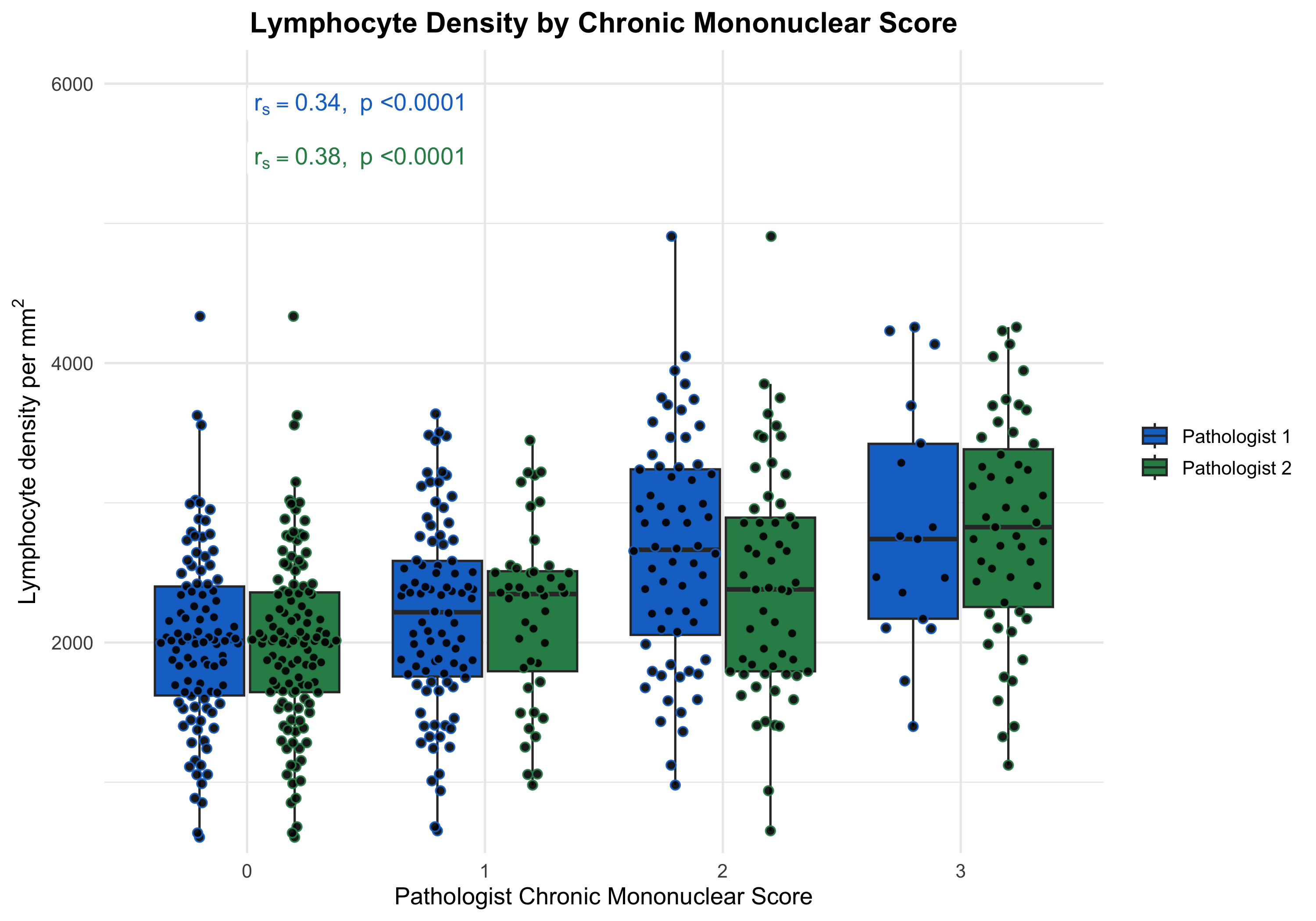

Supplementary Figure 3 – AI derived metrics compared in samples with different grades from manual reads (Universal Scoring Template). Crypt abscess was compared with pathologist scores for overall crypt inflammation (A) and crypt abscess score (B). Mucin depletion was compared with the mucin depletion pathologist’s score (C) in UC (n = 272).

**B)**

**C)**

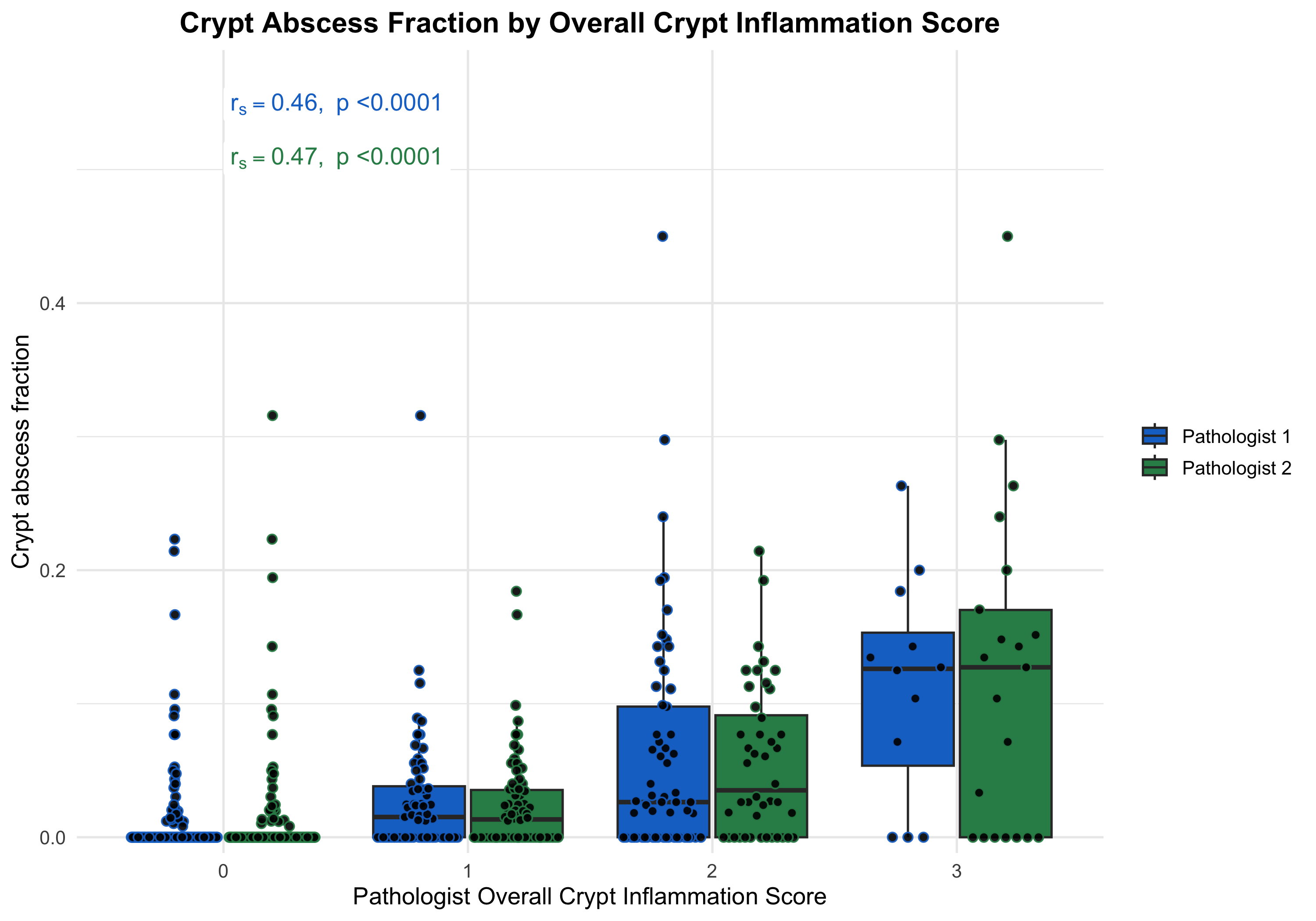

**A)**

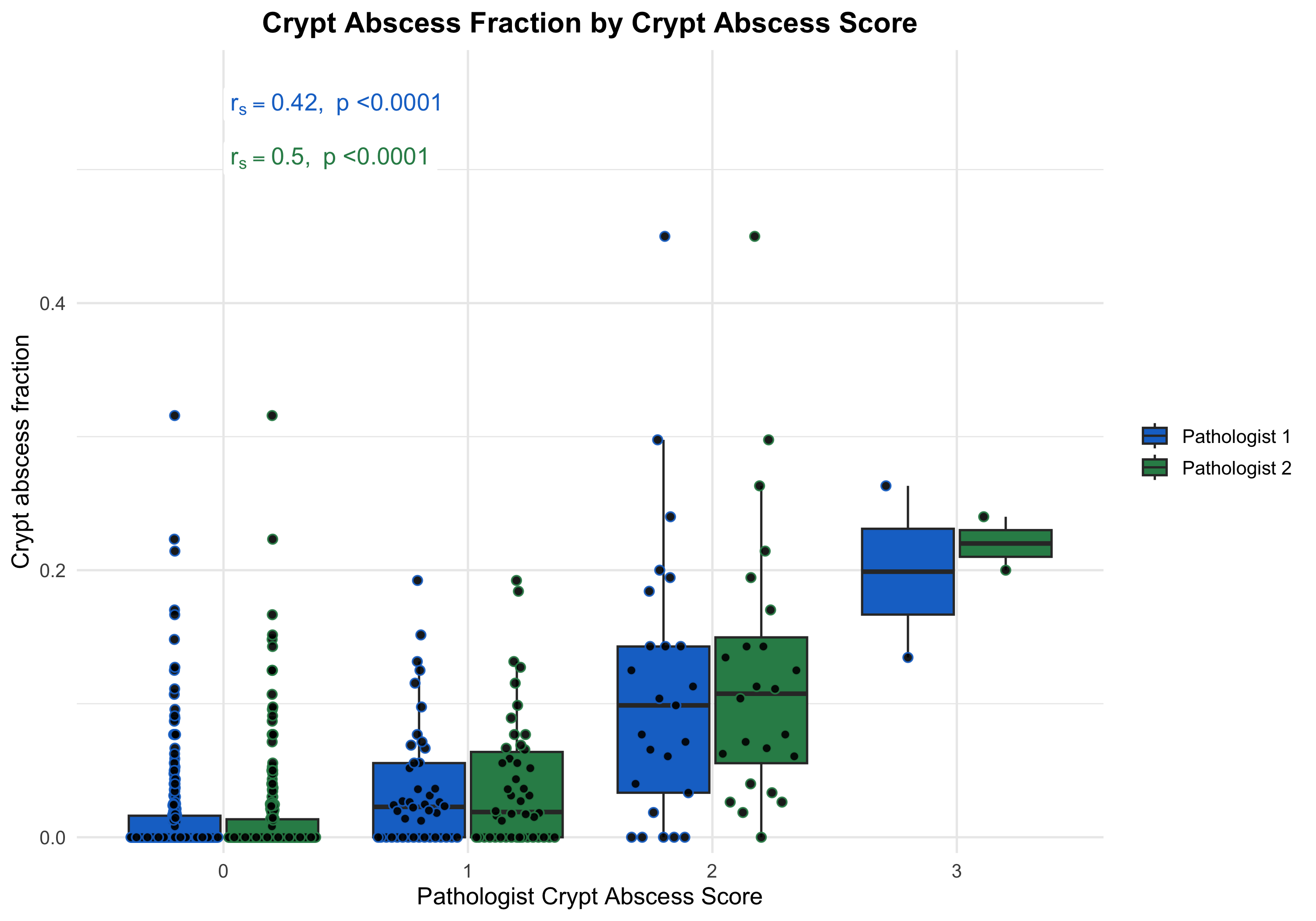

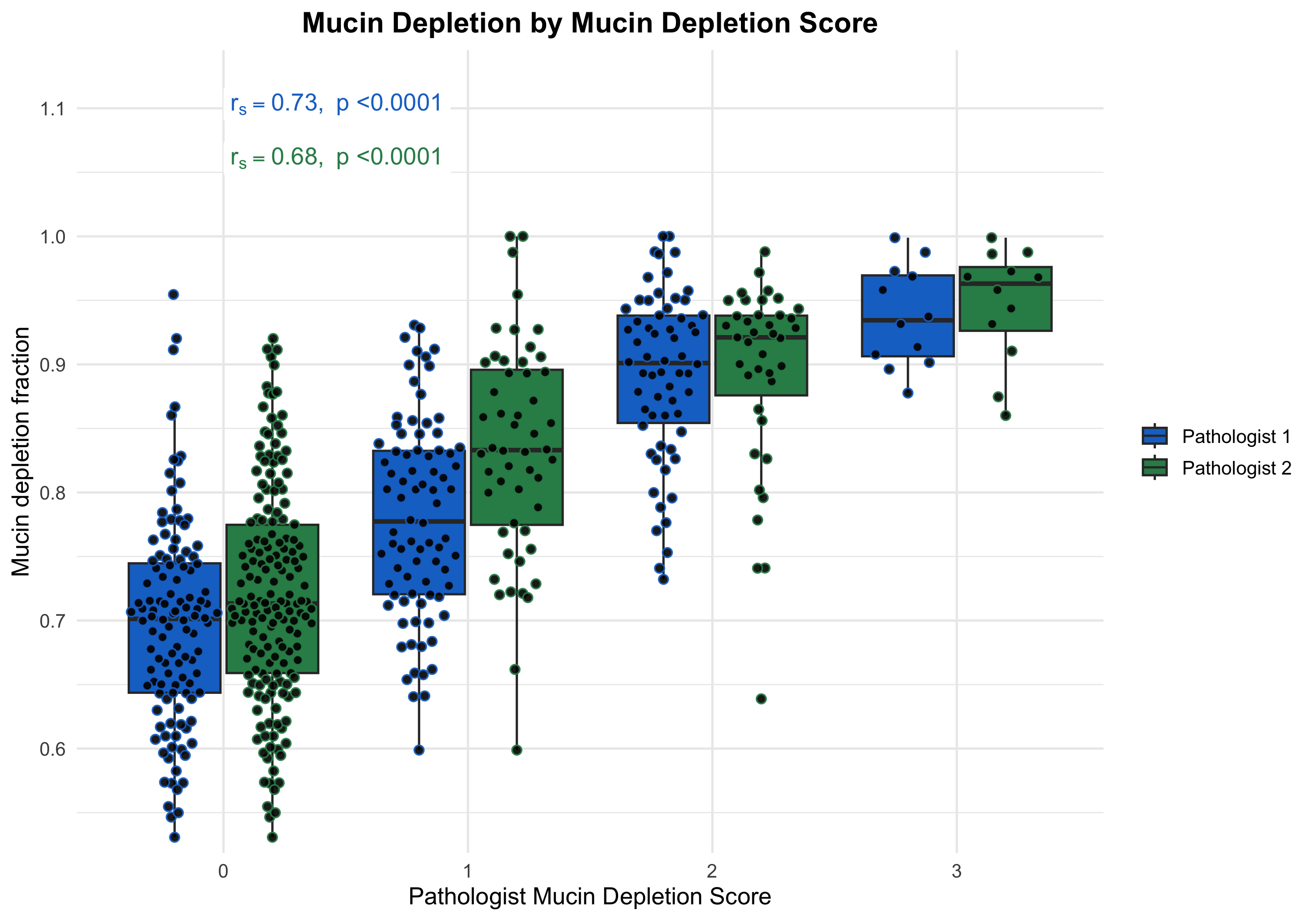

Supplementary Figure 4 – AI derived metrics compared in samples with different grades from manual reads (Universal Scoring Template). Crypt solidity was compared with pathologist scores for serrated architecture (A), crypt injury (B), epithelial injury (C), and architectural change (D) in UC (n = 272).

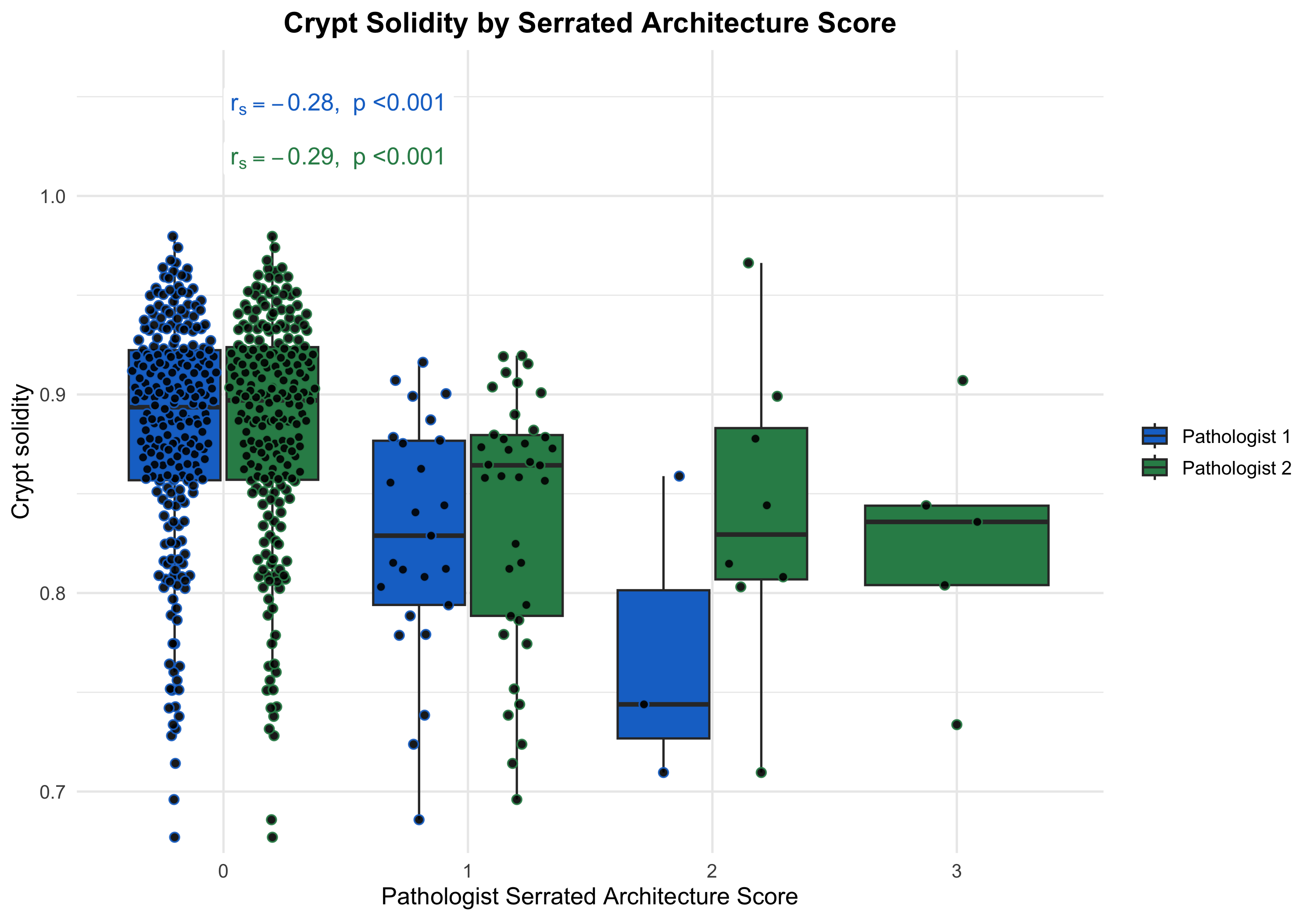

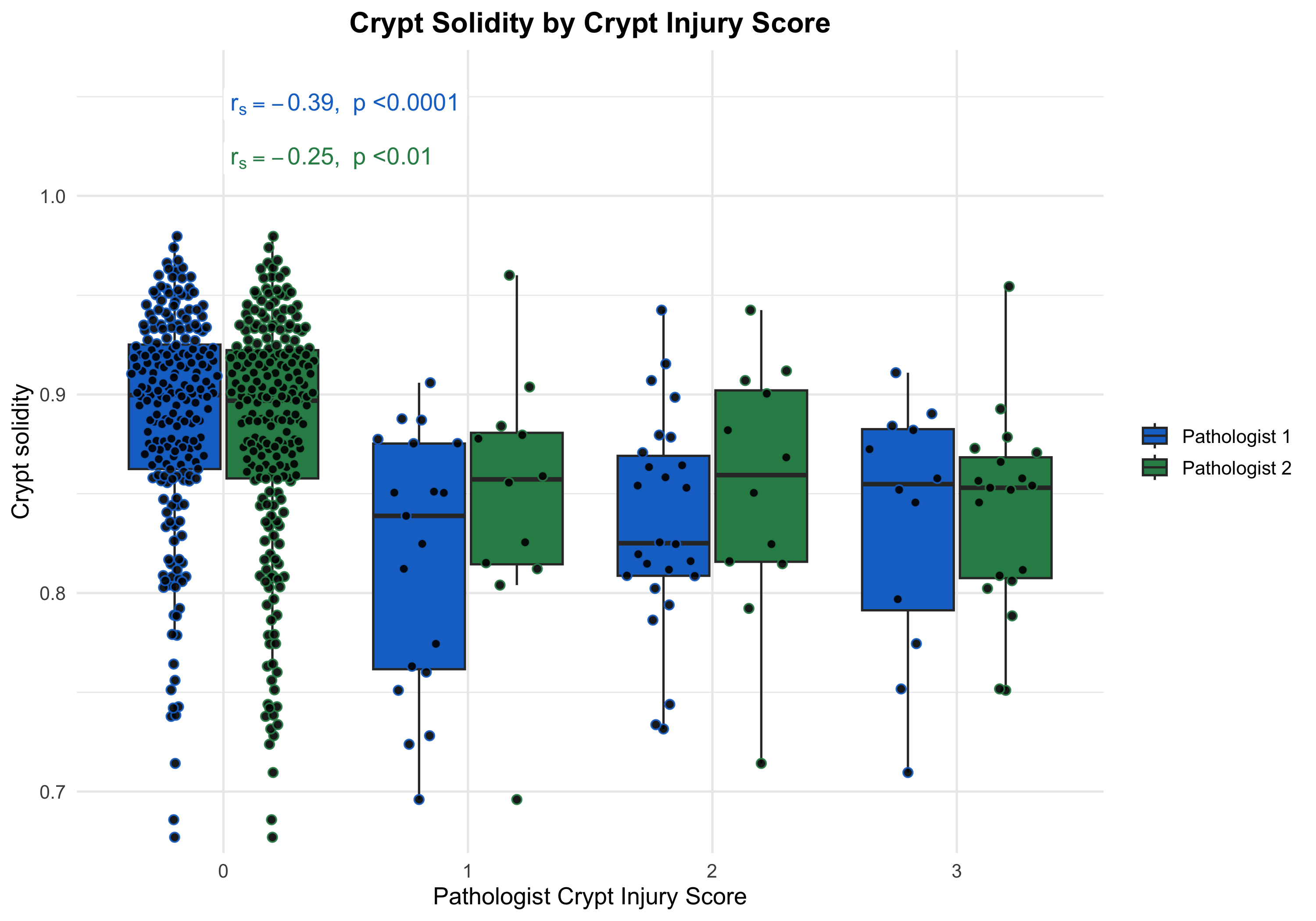

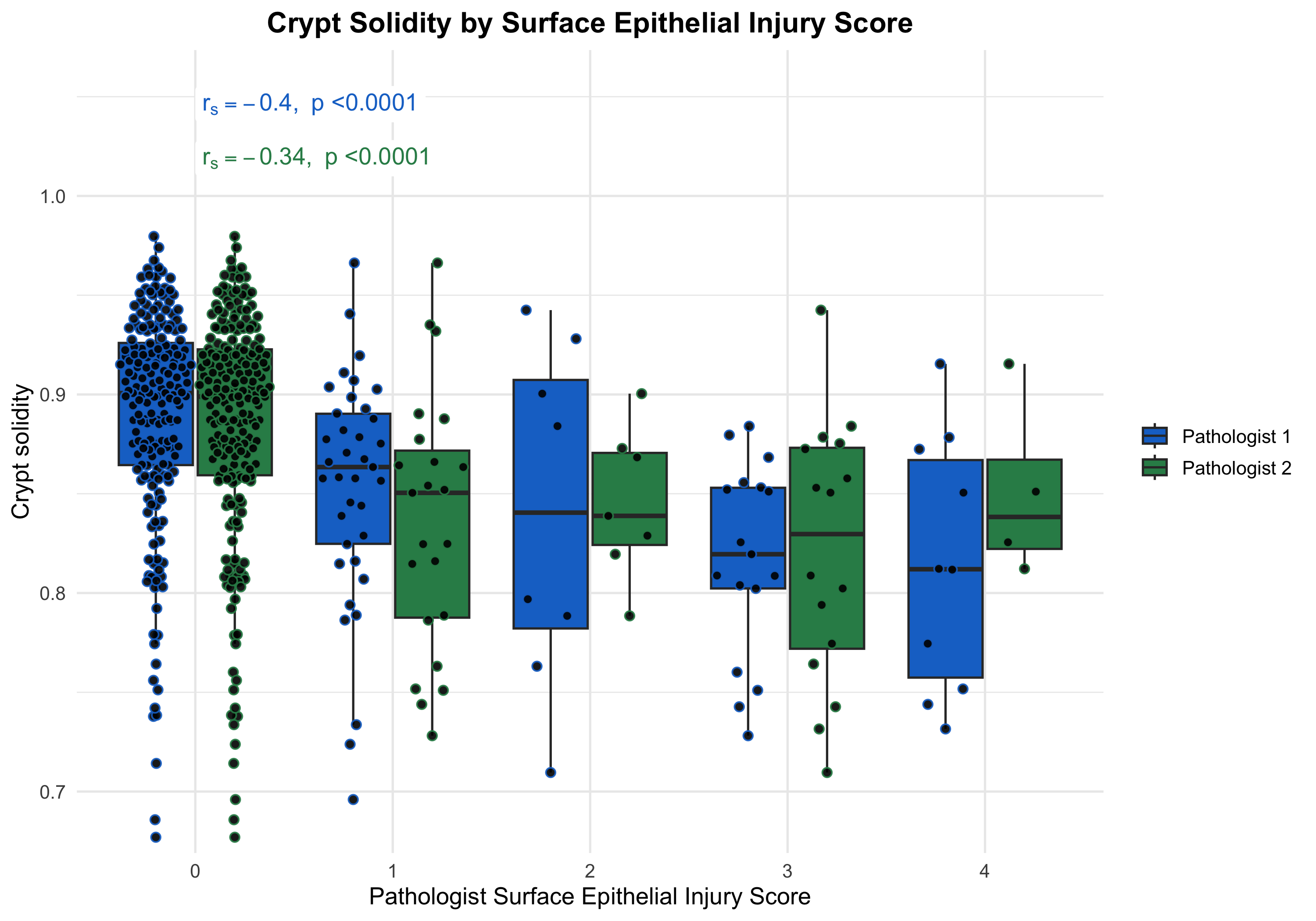

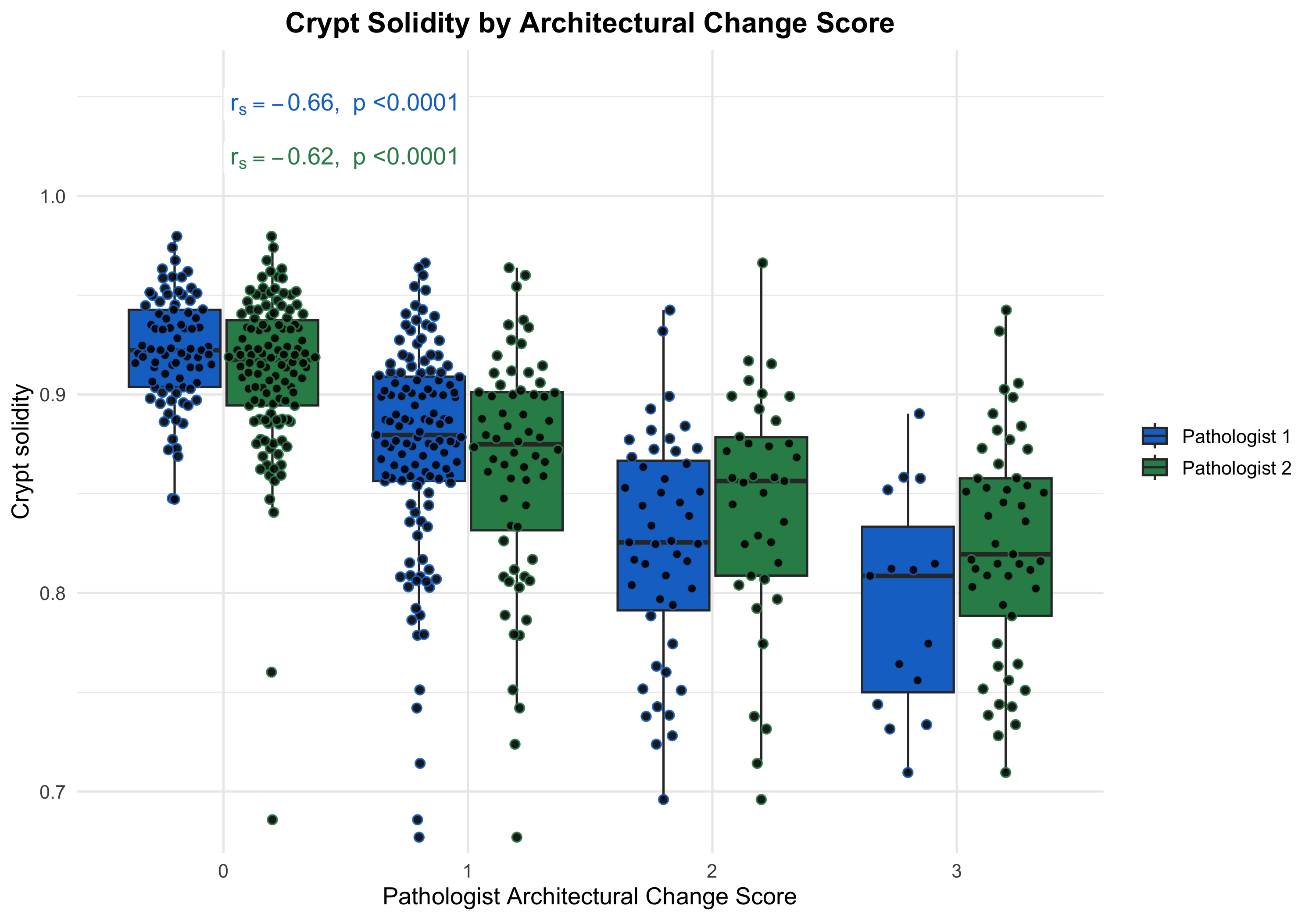

**B)**

**A)**

**D)**

**C)**

Supplementary Figure 5 – AI derived metrics compared in samples with different grades from manual reads (Universal Scoring Template). Crypt roughness was compared with pathologist scores for serrated architecture (A), crypt injury (B), epithelial injury (C), and architectural change (D) in UC (n = 272).

**B)**

**A)**

**D)**

**C)**

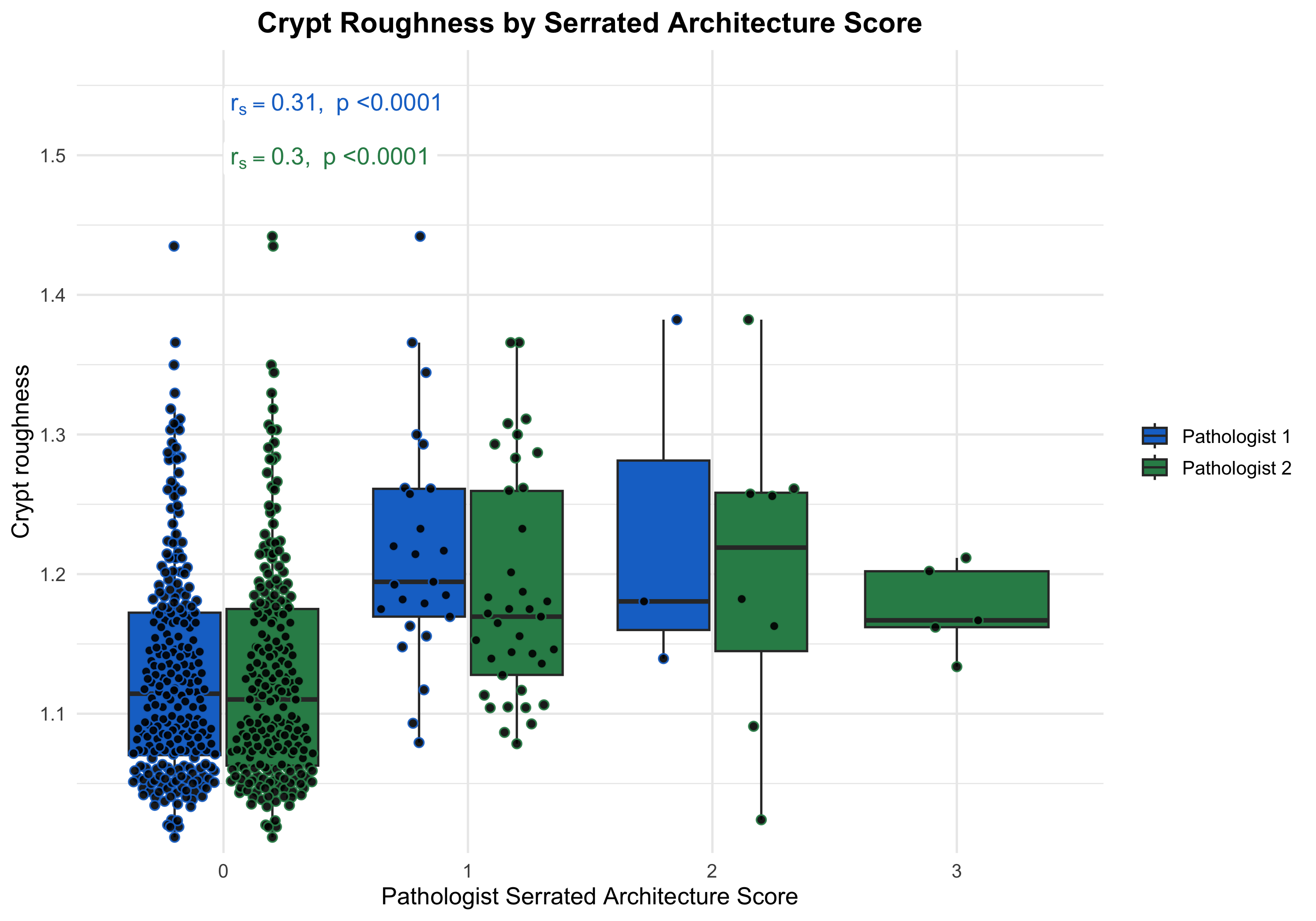

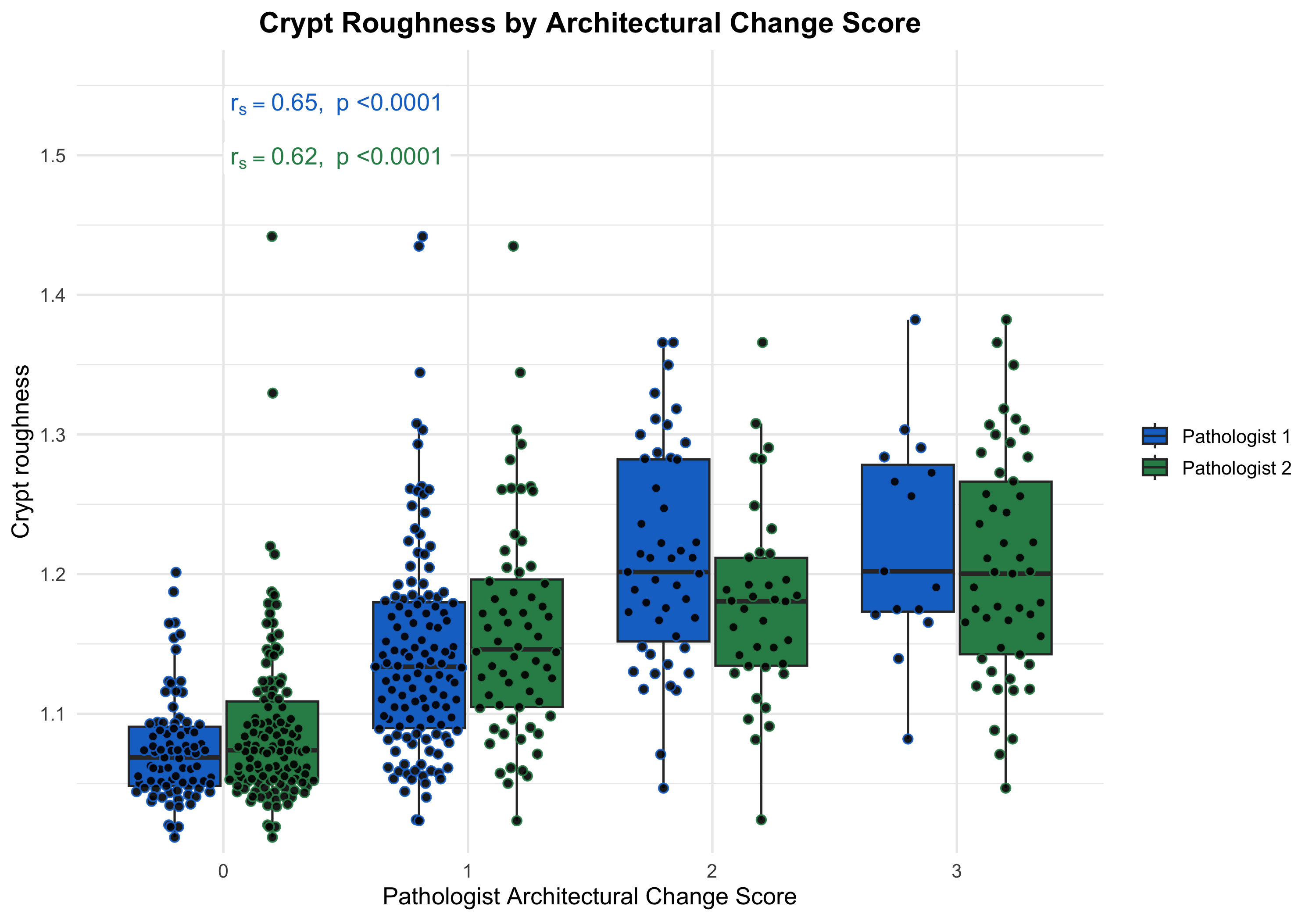

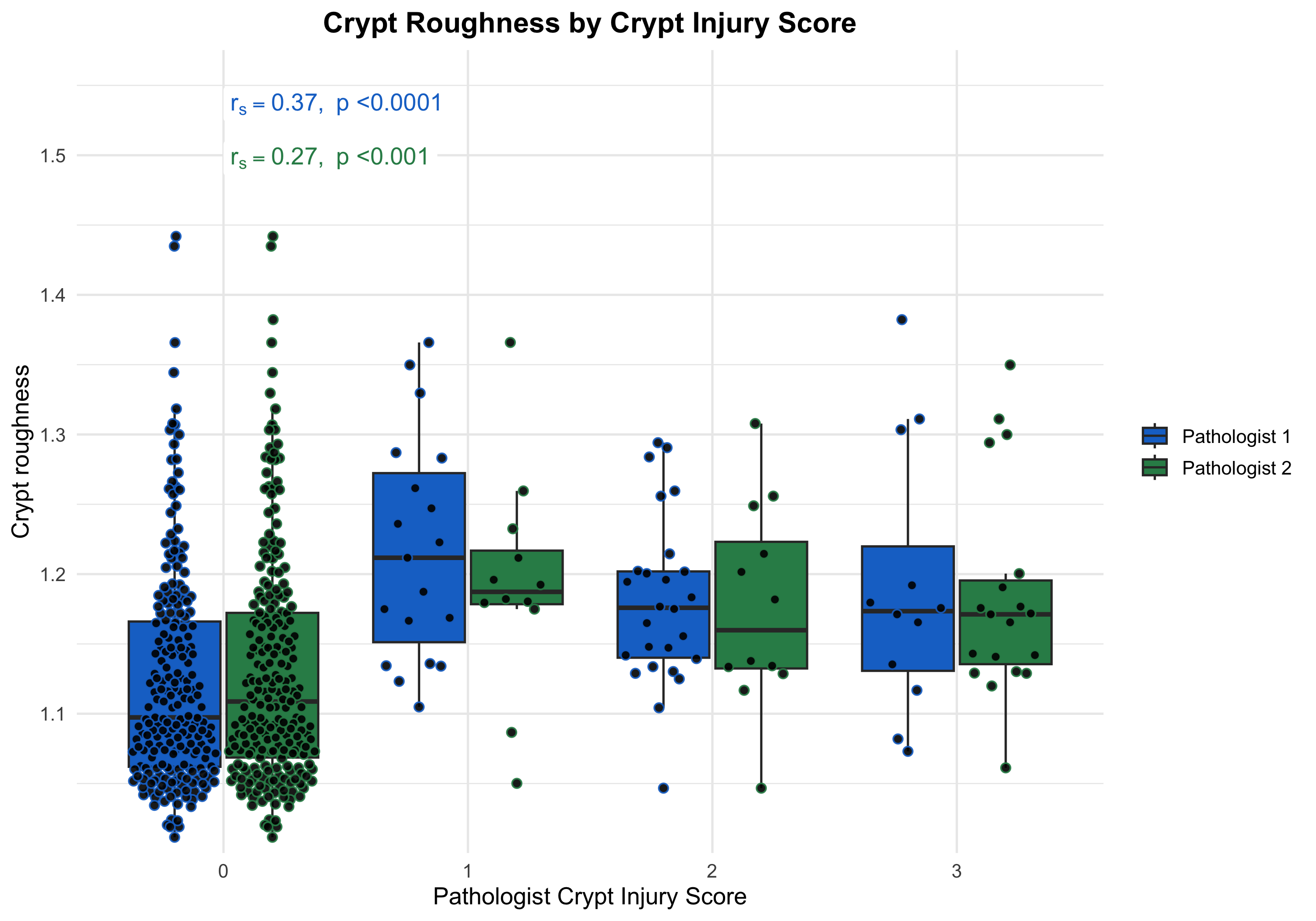

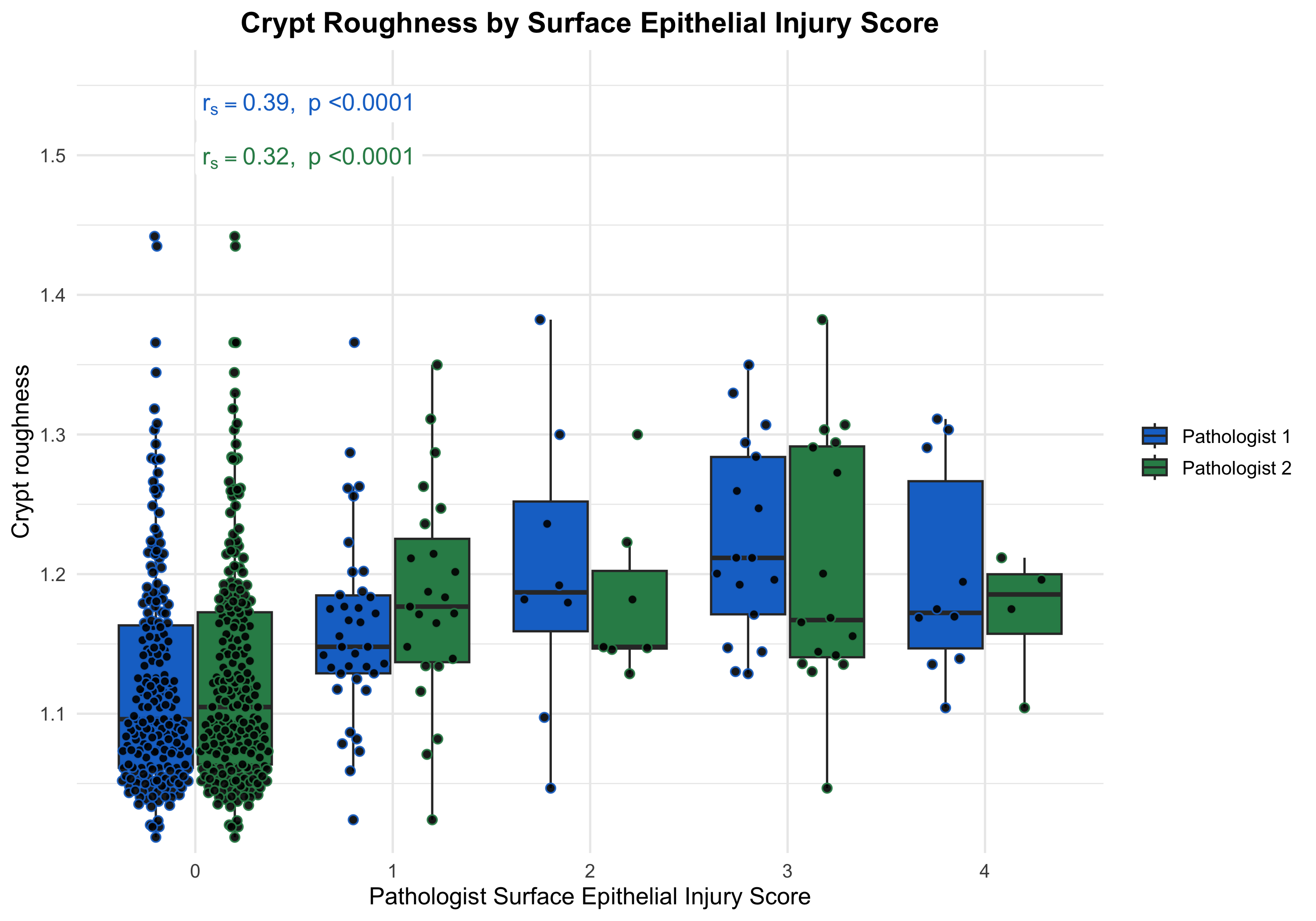

Supplementary Figure 6 – AI derived metrics compared in samples with different grades from manual reads (Universal Scoring Template). Crypt branches was compared with pathologist scores for serrated architecture (A), crypt injury (B), epithelial injury (C), and architectural change (D) in UC (n = 272).

**B)**

**A)**

**D)**

**C)**

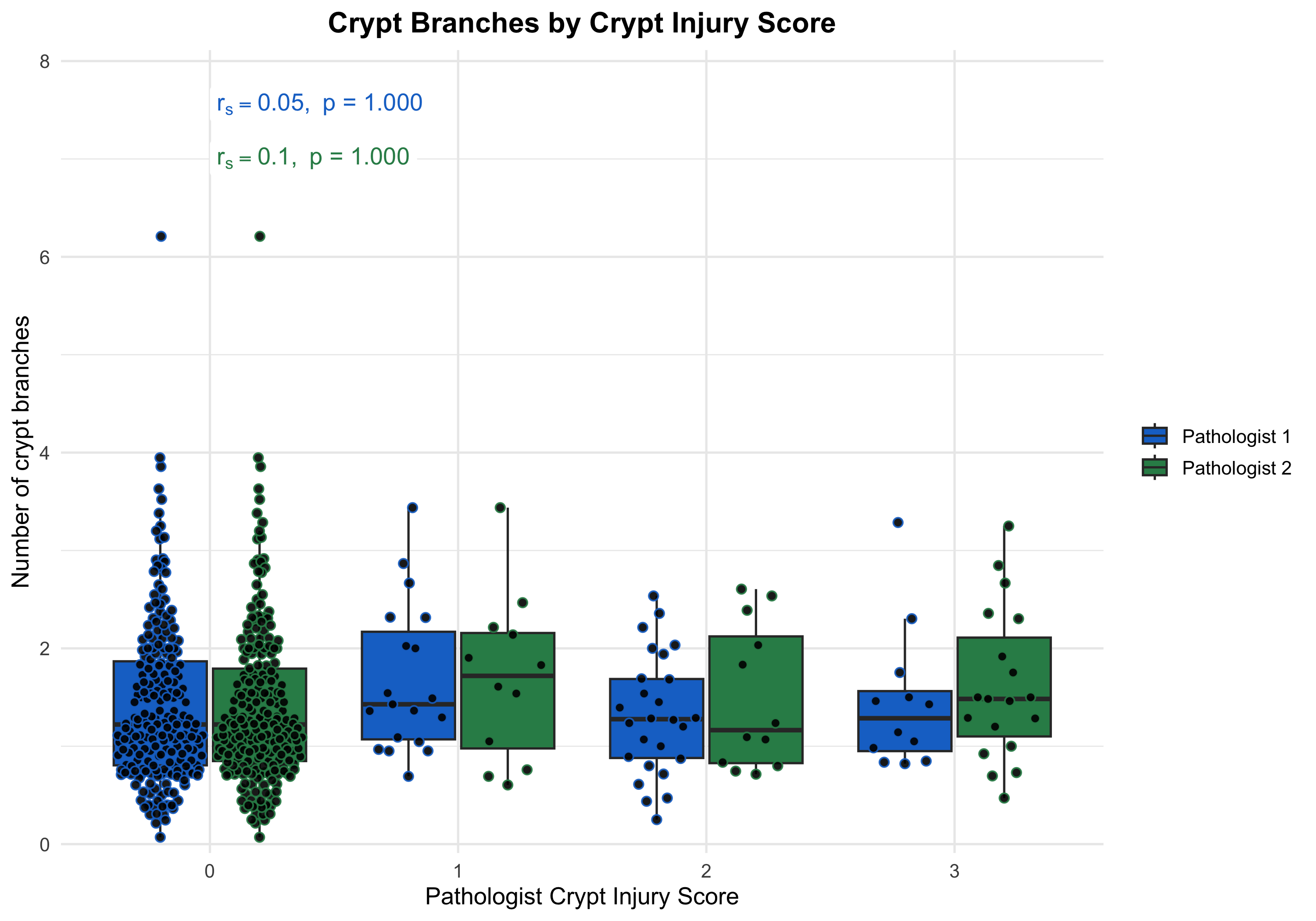

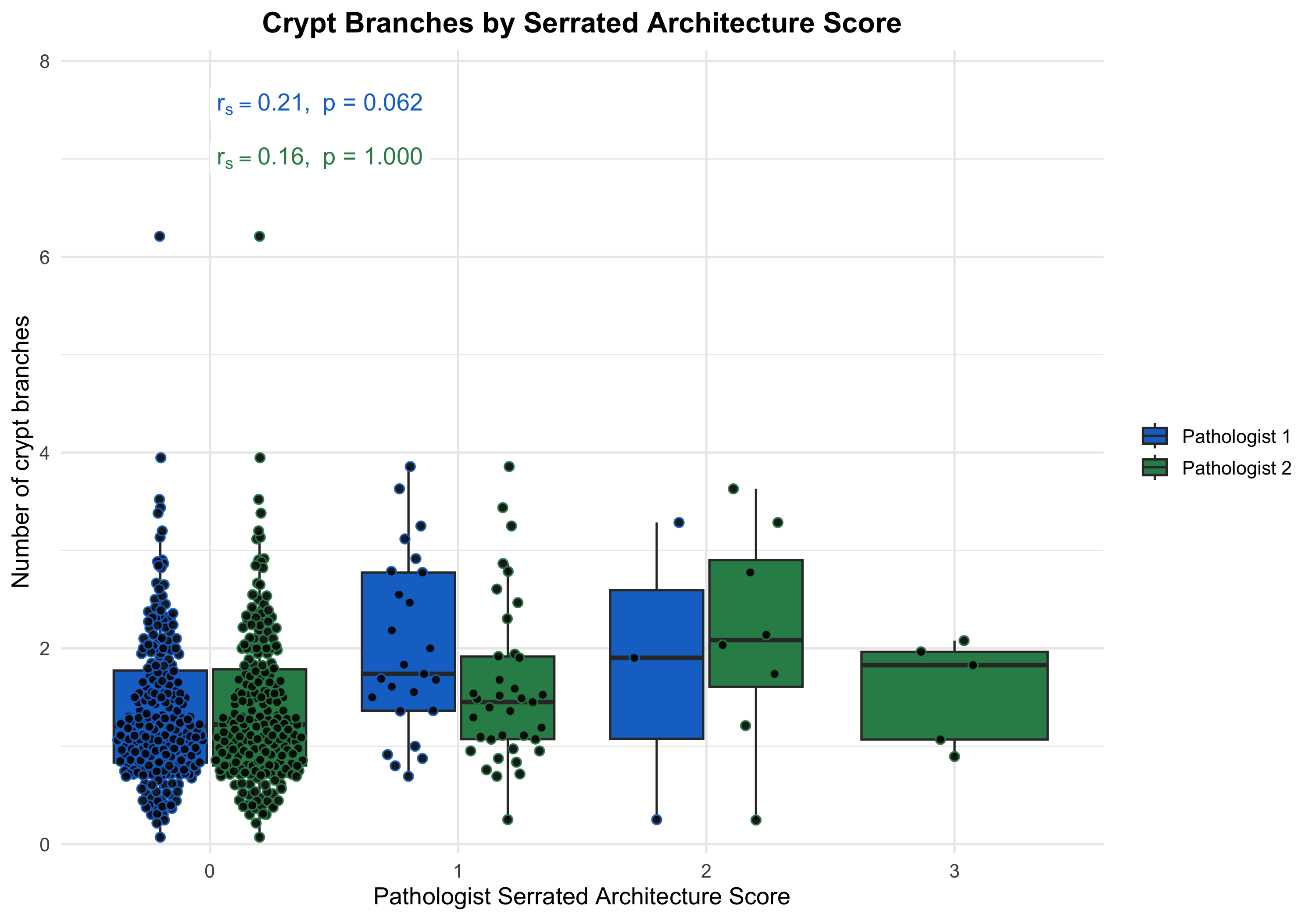

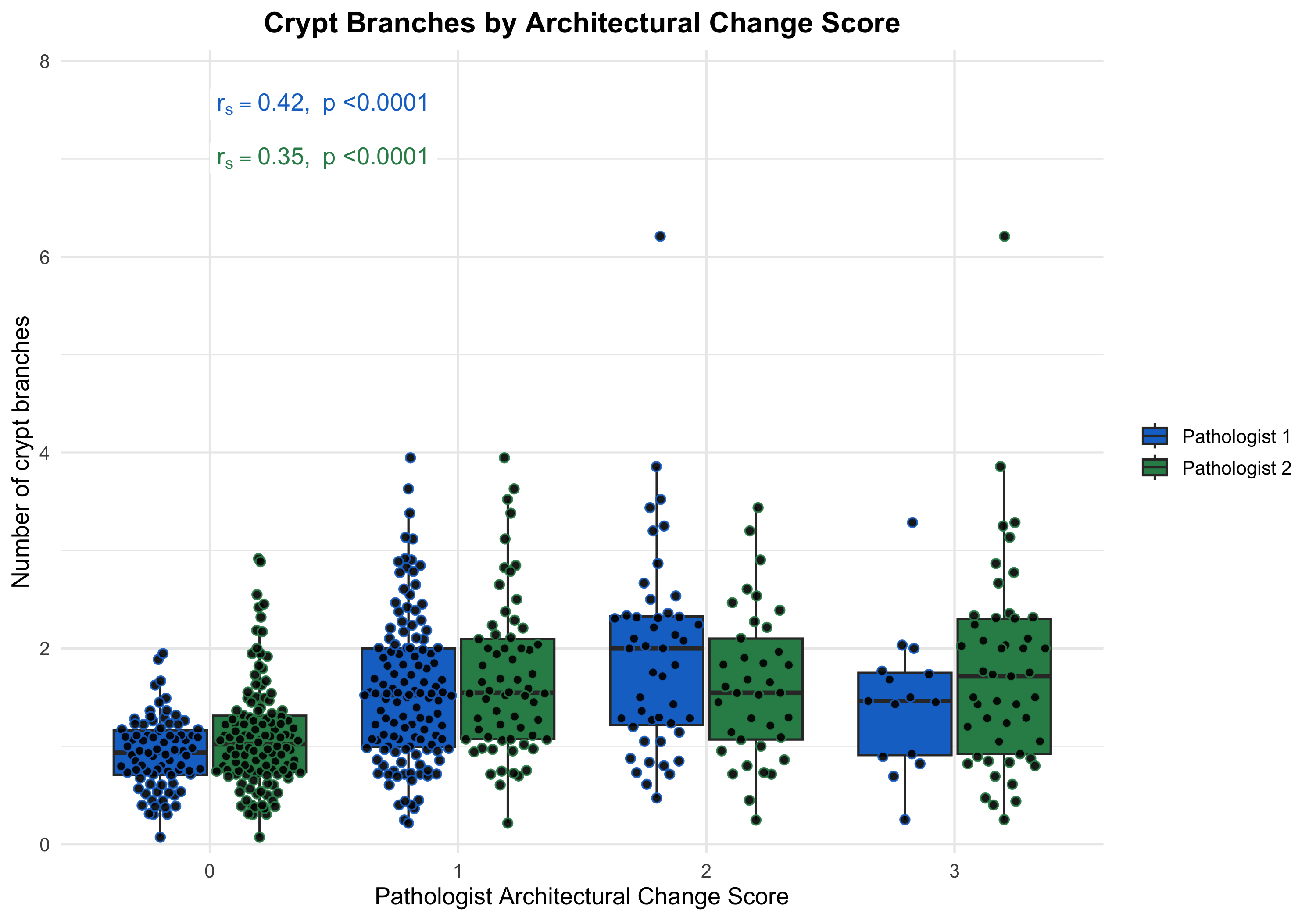

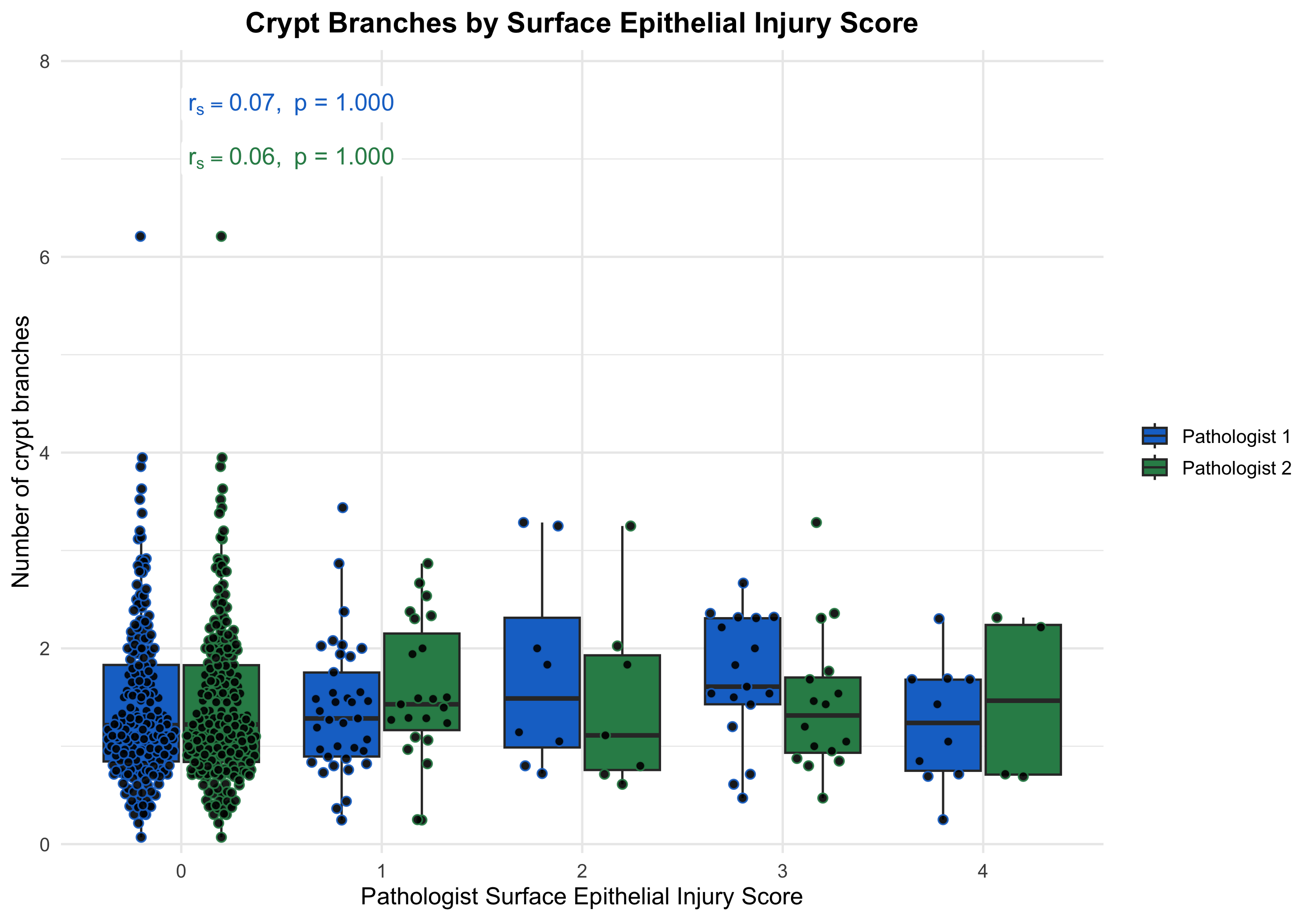

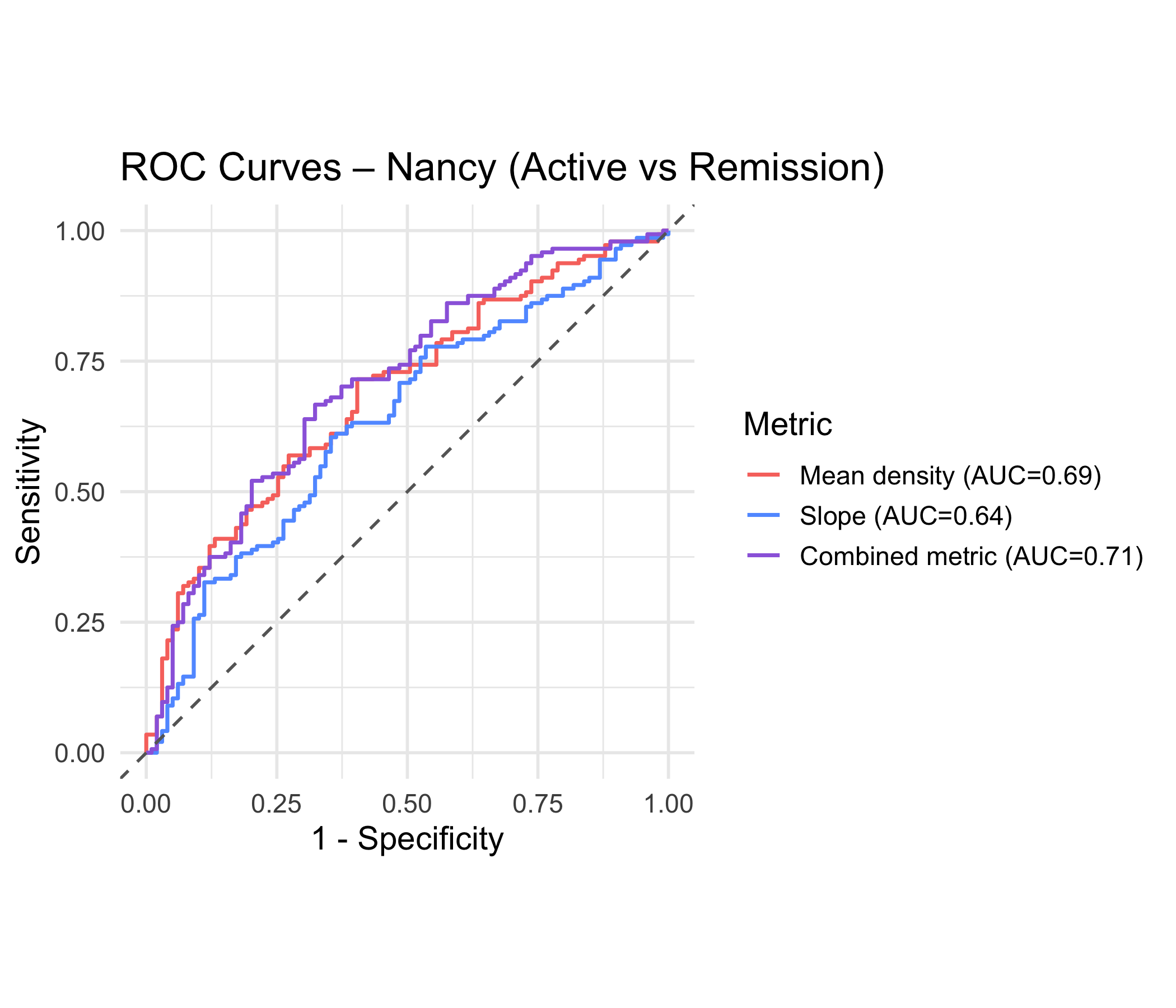

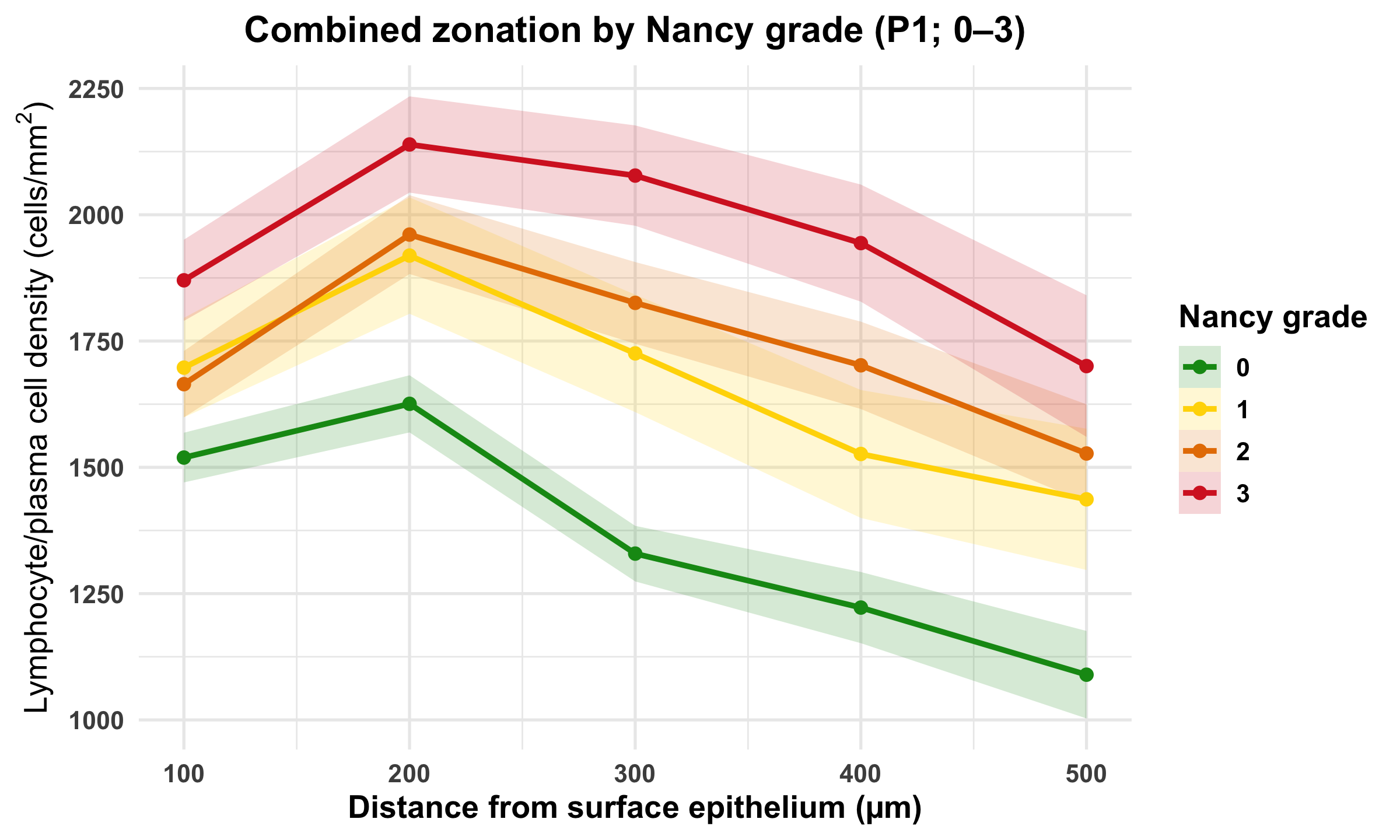

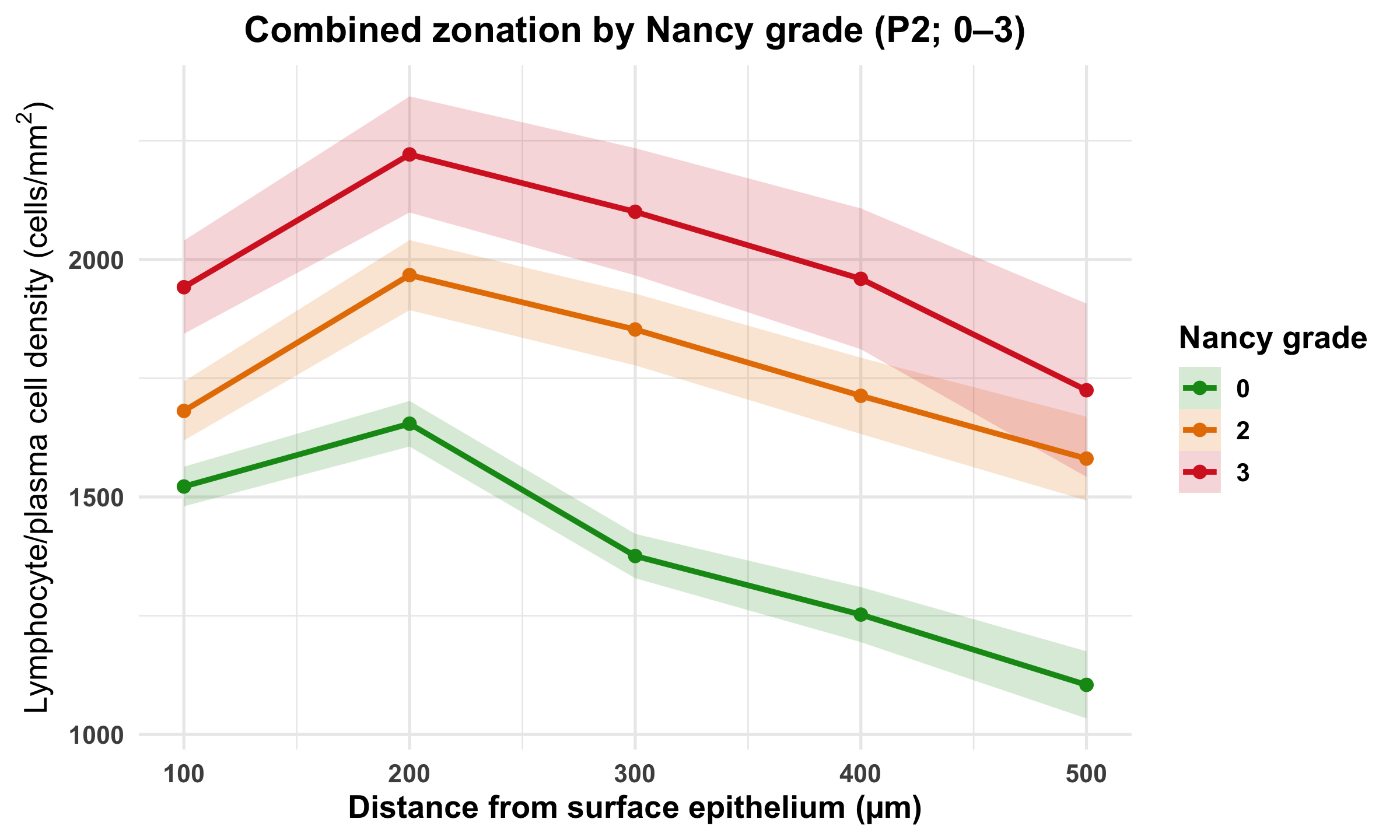

Supplementary Figure 7 – Combined lymphocyte/plasma cell zonation by Nancy grade and discrimination of active versus remission disease. (A-B) Combined lymphocyte/plasma cell density across five 100 μm zones from the surface epithelium toward the muscularis mucosa, stratified by Nancy grade (0–3) for P1 and P2, respectively. (C) ROC curves showing discrimination of active versus remission disease using mean density, profile AUC, and zonation slope.

**A)**

**B)**

**C)**
